# Long-Term Trajectories of Posttraumatic Stress Disorder Symptoms: A 20-Year Longitudinal Study of World Trade Center Responders

**DOI:** 10.1101/2024.03.25.24304851

**Authors:** Frank D. Mann, Monika A. Waszczuk, Sean A.P. Clouston, Scott Feltman, Camilo J. Ruggero, Brian P. Marx, Joseph E. Schwartz, Evelyn J. Bromet, Benjamin J. Luft, Roman Kotov

## Abstract

**Objective:** The present study examined the 20-year course of posttraumatic stress disorder (PTSD) in World Trade Center (WTC) responders to address four questions: (1) How stable are symptoms of PTSD? (2) What is the average symptom trajectory? (3) How much do responders differ from the average trend? (4) How quickly do PTSD symptoms improve or worsen?

**Methods:** Data include 81,298 observations from *n* = 12,822 responders, spanning from July 2002 to December 2022. Fourteen percent meet PTSD criteria. PTSD symptoms were measured using the PCL-17. Retest correlations were calculated to estimate stability, growth curve models to estimate individual trajectories, and Kaplan-Meier curves to estimate the rate of clinically significant change.

**Results:** Retest correlations were high overall (range =.49, .84), lower in PTSD cases (range =.21, .78), and decreased as a function of time between assessments. The best-fitting growth model represented trajectories continuously rather than multiple classes. Symptom burden peaked in 2011 and declined modestly by 2022 (Cohen’s d = -0.28 and -0.59 in all responders and PTSD cases, respectively). Median time before clinically significant improvement in responders with PTSD was 8.88 years (95% CI = 8.01, 9.79).

**Conclusions:** In the longest and largest study of PTSD symptoms tracked continuously since exposure, illness course was characterized to find that, while symptoms were highly stable in the short term, symptoms changed significantly over two decades. Most responders experienced clinical improvement after nine years, but 10% had poor course and should be the focus of public health efforts.

---

Post-traumatic stress disorder (PTSD) is a chronic and costly psychiatric condition that can persist decades after the triggering trauma [1]. World Trade Center (WTC) responders were exposed to extreme trauma during and after the 9/11 attacks. Similar to combat veterans, many WTC responders developed PTSD. For example, a decade after the 9/11 attacks, PTSD prevalence remained high (∼14%) among responders and other exposed populations [2, 3], comparable to veterans decades after the Yom Kippur War (8%) [4] and Vietnam War (8% - 14%) [5, 6]. While these similarities underscore the severity of WTC exposures and the chronicity of PTSD, existing long-term studies are limited by retrospective and infrequent measurement that often began years after traumatic exposure, precluding a comprehensive examination of symptom trajectories. We contribute to existing literature a detailed analysis of long-term symptom trajectories in one of the largest and longest studies of PTSD in a traumatized cohort to date, WTC responders.

Longitudinal studies have examined symptom course in different trauma-exposed populations, reporting average change across short and medium terms (1-10 years) [7-10]. These studies have shown that changes in PTSD symptoms vary greatly from person to person. Most exposed to trauma are resilient and never develop clinically elevated symptoms (stable low), but among those who do, symptom course varies from delayed onset (increasing) to chronic (stable high) and rapid improvement (decreasing) [4, 5, 11-13]. Yet, it is unknown whether these broad groupings adequately capture the long-term course of PTSD symptoms. To date, few studies have documented the long-term course of PTSD. One noteworthy exception [11] followed 1,353 U.S. veterans annually for 4.5 years, and used differences in timing of the precipitating trauma to model symptom trajectories over two decades following exposure. They found that symptoms increased slightly at first but improved significantly with accelerating pace over time, consistent with a pattern of remission. A replication is needed in another population and optimally with longer tracking of participants.

Temporal stability of PTSD symptoms can also be expressed as retest correlations. Short-term stability is very high with retest correlations of .71 to .77 over 2.5 to 4.5 years [14-16]. Long-term stability is lower but substantial, with .48 retest correlation in Israel veterans over 19 years [17]. However, it is unknown at what point symptoms become less stable and whether symptom stability differs in PTSD cases. Remission is another alternative description of course and has direct clinical implications. A meta-analysis found that remission rates varied wildly from 8% to 89% across naturalistic studies, and 44% of patients no longer met criteria for PTSD at follow-up, on average, 40 months after diagnosis [13]. Epidemiologic studies based on recall suggest that 50% of PTSD cases remit within 24 months and 77% within 10 years [12]. However, long-term probability of remission is unknown, and the likelihood of symptoms worsening is understudied.

The present study analyzed 81,298 observations from 12,822 individuals, starting immediately after the exposure concluded and spanning more than 20 years of data collection, including as many as 17 follow-up assessments per individual. Specifically, we sought to answer four questions: (1) How stable are symptoms of PTSD? (2) What is the typical direction, shape, and rate of change? (3) What are atypical trajectories? (4) How quickly do PTSD symptoms improve or worsen?

## Methods

### Sample

Participants are members of WTC Health Program Long Island Clinical Center of Excellence [18, 19]. They are eligible for the program because of a qualifying exposure during response to 9/11 attacks. Members are invited for annual monitoring visits for WTC-related disorders. If PTSD is detected, members are offered free treatment, and 76% of cases have received evidence-based mental health treatment [20]. The response efforts officially concluded in June 2002, when the fires from the WTC were all finally extinguished. The program began monitoring responders in July 2002, and has continued annual follow-up evaluations while maintaining open enrollment.

### Measures

#### Self-Reported Symptoms

PTSD symptoms were assessed using the trauma-specific version of the PTSD Checklist (PCL-17) [21], adapted for the WTC disaster. Participants rate past-month DSM-IV symptoms from 1 (not at all) to 5 (extremely), total scores range = 17 to 85.

#### Diagnostic Interviews

Diagnosis of PTSD was assigned by trained clinicians using the Diagnostic Interview Schedule (DIS) [22] or the Structured Clinical Interview for DSM-IV (SCID) [23]. Participants were interviewed ∼12 years after 9/11 (mean = 12.20, range = 10.33–15.07), and inter-rater agreement was high (kappa > 0.82) for a subset of independently rated audio-taped interviews [20]. In total, 7,298 responders (56.9%) completed the DIS or SCID, resulting in 1,007 lifetime cases of WTC-related PTSD (13.8% of interviewed responders). There were no significant differences in demographic characteristics by interview status [24], though PCL scores at enrollment were slightly higher in those who did not complete a diagnostic interview (*Hedges g* = 0.03, *p* < 0.001).

### Data Analysis

All analyses were conducted separately for the full sample (*n* = 12,822) and for individuals with diagnosed PTSD (*n* = 1,007). First, descriptive statistics and internal consistency were calculated for PCL total scores at each year and visit, and individual trajectories were visualized (Figures S1 and S2). Second, temporal stability of the PCL was calculated as Pearson’s retest correlations across 1-year to 20-year time-lags. Correlations were aggregated for each lag and standard deviations calculated using meta-analytic methods [25]. Third, mixed effects models were used to estimate individual PCL trajectories since 9/11, with unconstrained correlations among random effects and time centered 10 years after the attacks.

In total, nine mixed models were estimated, beginning with a “null” intercept-only model that allowed only individual differences in average levels, and progressing to models that allowed for linear and non-linear changes (see Supplemental Methods). To address deviation from normality, linear mixed effects models (with an identity link and Gaussian distribution) were compared to generalized mixed effect models (with a log link and Gamma distribution). To assess whether individual trajectories form discrete groups, latent class growth models (LCGMs) and growth mixture models (GMMs) were fit to the data (see Table S4). Information criteria (*AIC, BIC, SSBIC*) were used to guide the selection of a preferred model, along with the interpretability of parameter estimates and ability to accurately predict mean change and subject-specific trajectories. The model estimator (maximum likelihood) and optimization algorithm (bound optimization by quadratic approximation) were held constant across mixed models, LCGMs, and GMMs to ensure these features had no impact on the likelihood function and comparison of information criteria. Results were also compared across different R packages (“nlme”, “lme4”, “lcmm”) to ensure consistency of findings across analytic routines. Finally, to assess the amount of time until symptoms improved or worsened, clinically significant change was defined as a 10-point increase or decrease on the PCL [26]. This threshold was identified for raw scores—rather than predicted trajectories—and therefore Kaplan-Meier analyses examined time since first visit until a 10-point change was observed (increase or decrease, not mutually exclusive).

## Results

PTSD symptoms were assessed between 07/19/2002 and 12/29/2022 for 12,822 responders, with visits approximately one year apart (mean = 1.27 years, SD = 1.29 years). Responders complete 6.3 visits on average (SD = 4.20), resulting in 81,298 observations across 20.45 years. Table 1 reports observations per year (Table S1 reports observations per visit). Monitoring begun with 300 visits in 2002, which steadily increased to 9,008 in 2022 (Table 1). PCL consistently showed high internal consistency (Cronbach’s α = .94 to .96 and μ_T_ = .95 to .97). Based on a PCL-17 total score of ≥44 [27], the prevalence of clinical PTSD symptoms ranged from 8% to 15% between 2002 to 2022 (Figure 1A). Retest correlations were very high over one year (*r* > .80) but decreased steadily (b = -0.018 [-0.019, -0.018], *p* < 0.001) with increasing time-lags to *r* = .49 over 20 years (Figure 1B). Cases showed a more rapid decrease in retest correlations (*b* = -0.029 [-0.031, 0.027], *p* < 0.001) down to *r* = .21 over 20 years.

**Table 1A.** Descriptive Statistics and Internal Consistency of PCL Scores By Year of Data Collection

| Year | Descriptive Statistics |  |  |  |  |  | Internal Consistency |  |  |
| --- | --- | --- | --- | --- | --- | --- | --- | --- | --- |
| | n | mean | median | SD | min | max | $\alpha$ | $\omega_T$ | $\omega_H$ |
| 2002 | 300 | 29.27 | 24.50 | 12.57 | 17.00 | 74.00 | 0.94 | 0.96 | 0.79 |
| 2003 | 875 | 27.88 | 24.00 | 11.41 | 17.00 | 78.00 | 0.94 | 0.95 | 0.81 |
| 2004 | 703 | 26.43 | 22.31 | 11.24 | 17.00 | 81.00 | 0.95 | 0.96 | 0.80 |
| 2005 | 766 | 26.97 | 22.31 | 12.25 | 17.00 | 85.00 | 0.95 | 0.96 | 0.82 |
| 2006 | 1003 | 29.16 | 24.00 | 13.70 | 17.00 | 85.00 | 0.96 | 0.96 | 0.84 |
| 2007 | 1616 | 28.57 | 24.00 | 13.16 | 17.00 | 85.00 | 0.95 | 0.96 | 0.82 |
| 2008 | 1552 | 28.71 | 24.00 | 13.59 | 17.00 | 83.00 | 0.96 | 0.96 | 0.85 |
| 2009 | 2750 | 27.98 | 23.00 | 13.46 | 17.00 | 85.00 | 0.96 | 0.96 | 0.84 |
| 2010 | 3112 | 28.33 | 23.00 | 13.80 | 17.00 | 85.00 | 0.96 | 0.97 | 0.86 |
| 2011 | 3116 | 29.70 | 23.38 | 14.77 | 17.00 | 85.00 | 0.96 | 0.97 | 0.85 |
| 2012 | 3341 | 28.78 | 23.00 | 13.84 | 17.00 | 85.00 | 0.96 | 0.96 | 0.84 |
| 2013 | 3811 | 28.56 | 23.00 | 13.65 | 17.00 | 85.00 | 0.96 | 0.96 | 0.85 |
| 2014 | 4438 | 28.33 | 23.00 | 13.67 | 17.00 | 85.00 | 0.96 | 0.97 | 0.87 |
| 2015 | 4017 | 28.20 | 22.00 | 13.68 | 17.00 | 84.00 | 0.96 | 0.97 | 0.87 |
| 2016 | 5137 | 28.42 | 23.00 | 13.61 | 17.00 | 85.00 | 0.96 | 0.97 | 0.86 |
| 2017 | 5515 | 28.47 | 23.00 | 13.65 | 17.00 | 84.00 | 0.96 | 0.97 | 0.86 |
| 2018 | 6521 | 27.80 | 22.00 | 13.27 | 17.00 | 85.00 | 0.96 | 0.97 | 0.86 |
| 2019 | 7622 | 27.67 | 22.00 | 13.13 | 17.00 | 85.00 | 0.96 | 0.96 | 0.86 |
| 2020 | 7039 | 26.84 | 22.00 | 12.65 | 17.00 | 85.00 | 0.96 | 0.96 | 0.85 |
| 2021 | 9056 | 26.51 | 21.00 | 12.38 | 17.00 | 85.00 | 0.96 | 0.96 | 0.84 |
| 2022 | 9008 | 26.04 | 21.00 | 12.01 | 17.00 | 85.00 | 0.96 | 0.96 | 0.85 |

**Table 1B.** Descriptive Statistics and Internal Consistency of PCL Scores By Year of Data Collection for WTC Responders with a Lifetime Diagnosis of PTSD

| Year | n | mean | median | SD | min | max | $\alpha$ | $\omega_T$ | $\omega_H$ |
| --- | --- | --- | --- | --- | --- | --- | --- | --- | --- |
| 2002 | 38.00 | 41.84 | 41.50 | 15.03 | 18.00 | 72.25 | 0.94 | 0.96 | 0.72 |
| 2003 | 90.00 | 38.22 | 35.53 | 13.05 | 18.00 | 78.00 | 0.92 | 0.94 | 0.74 |
| 2004 | 72.00 | 40.86 | 39.00 | 15.50 | 17.00 | 80.00 | 0.94 | 0.96 | 0.87 |
| 2005 | 86.00 | 43.07 | 44.50 | 17.68 | 18.00 | 85.00 | 0.96 | 0.97 | 0.84 |
| 2006 | 129.00 | 45.63 | 45.00 | 16.03 | 17.00 | 85.00 | 0.94 | 0.96 | 0.78 |
| 2007 | 197.00 | 45.69 | 46.00 | 16.29 | 17.00 | 85.00 | 0.94 | 0.95 | 0.77 |
| 2008 | 192.00 | 47.07 | 45.50 | 16.15 | 17.00 | 83.00 | 0.94 | 0.95 | 0.81 |
| 2009 | 361.00 | 46.10 | 45.69 | 16.57 | 17.00 | 85.00 | 0.95 | 0.96 | 0.82 |
| 2010 | 406.00 | 47.60 | 47.80 | 16.55 | 17.00 | 85.00 | 0.94 | 0.96 | 0.82 |
| 2011 | 439.00 | 49.59 | 49.00 | 16.08 | 17.00 | 85.00 | 0.94 | 0.95 | 0.83 |
| 2012 | 467.00 | 47.37 | 46.00 | 15.44 | 17.00 | 85.00 | 0.93 | 0.95 | 0.80 |
| 2013 | 516.00 | 46.21 | 45.00 | 15.52 | 17.00 | 85.00 | 0.94 | 0.95 | 0.79 |
| 2014 | 564.00 | 45.99 | 45.00 | 15.91 | 17.00 | 85.00 | 0.94 | 0.95 | 0.82 |
| 2015 | 487.00 | 45.92 | 45.00 | 15.40 | 17.00 | 81.00 | 0.94 | 0.95 | 0.78 |
| 2016 | 602.00 | 45.59 | 46.00 | 15.57 | 17.00 | 85.00 | 0.94 | 0.95 | 0.81 |
| 2017 | 649.00 | 46.57 | 46.14 | 15.20 | 17.00 | 83.00 | 0.94 | 0.95 | 0.80 |
| 2018 | 713.00 | 46.05 | 46.00 | 15.10 | 17.00 | 85.00 | 0.93 | 0.95 | 0.76 |
| 2019 | 736.00 | 45.65 | 45.00 | 15.08 | 17.00 | 85.00 | 0.93 | 0.95 | 0.77 |
| 2020 | 603.00 | 43.18 | 42.00 | 15.55 | 17.00 | 85.00 | 0.94 | 0.95 | 0.81 |
| 2021 | 754.00 | 41.46 | 40.00 | 15.46 | 17.00 | 85.00 | 0.94 | 0.95 | 0.81 |
| 2022 | 698.00 | 40.33 | 39.00 | 14.75 | 17.00 | 85.00 | 0.94 | 0.95 | 0.79 |

**Notes.** n = sample size. SD = standard deviation. min. = minimum observed scores. max. = maximum observed score. $\alpha$ = Cronbach's alpha. $\omega_T$ = Omega total. $\omega_H$ = Omega hierarchical.

**Figure 1.**
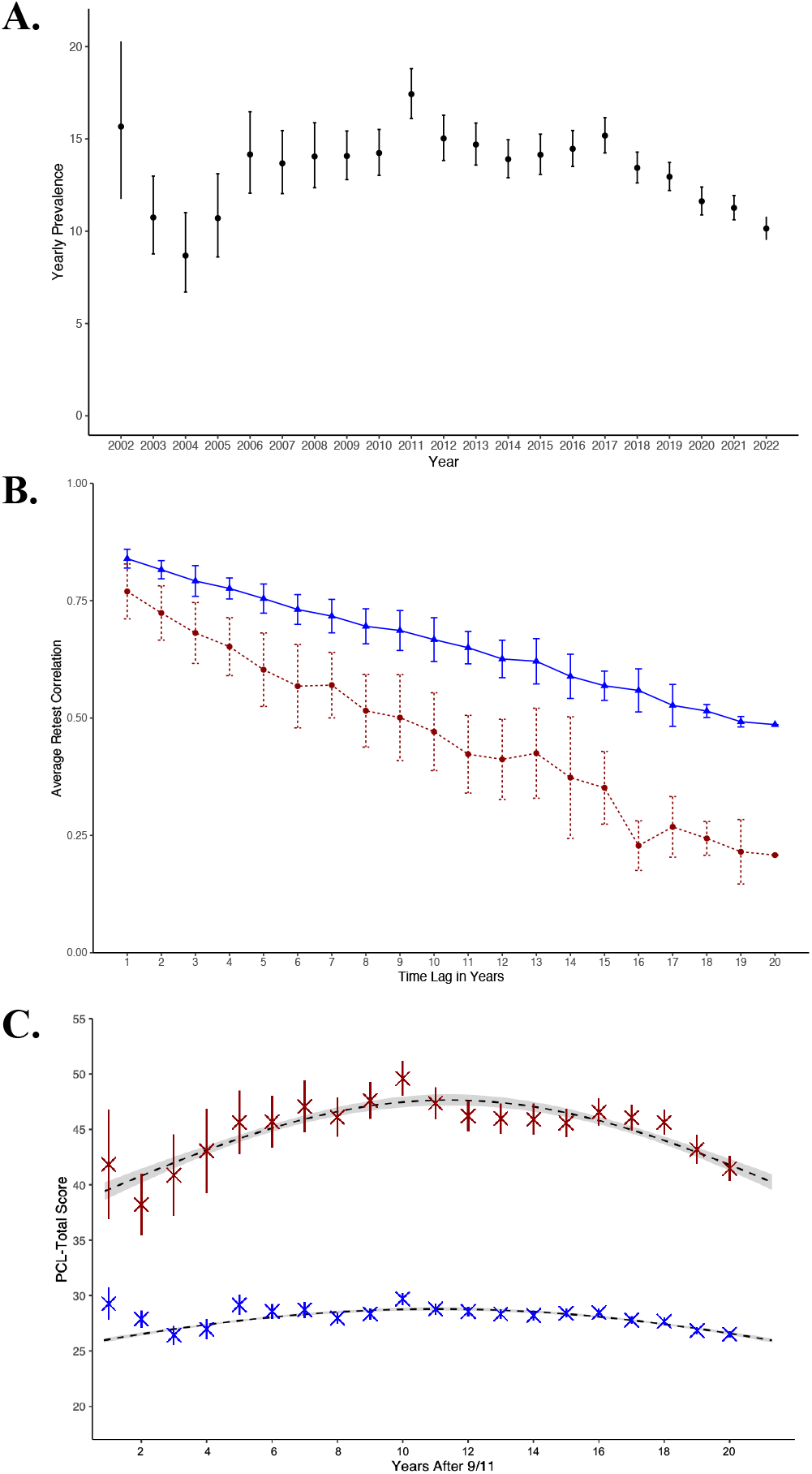
Annual Prevalence of Clinically Elevated Symptoms, Rank-order Stability, and Mean Predicted Trajectory of PTSD Symptoms **Notes. A**. Annual prevalence of clinical symptoms (PCL ≥ 44) with bars denoting 95% confidence intervals. **B**. Variogram depicted mean Pearson’s correlations (y-axis) across increasing time lags in years (x-axis) for the full sample (blue triangles) and for PTSD cases (red circles) with error bars denoting +/- 1 standard deviation. Full correlation matrices are given in Figures S3 and S4. **C**. Predicted mean trajectories (dashed lines) for the full sample and PTSD cases based on generalized mixed effects models with a Gamma distribution and log link with 95% confidence bands shaded in gray. “X” indicates annual observed means for the full sample (blue) and for PTSD cases (red) +/- 2 times the standard error of the mean. Plots further stratified on diagnosis of PTSD versus no PTSD are reported in Figures S7-S9.

### Longitudinal Model Selection

Comparison of mixed effects models showed that generalized mixed effects models fit the data much better than their parametric counterparts (information criteria improved >6000; (Tables S2-S3). For both parametric and generalized models the three with the lowest information criteria included linear, quadratic, and in some instances cubic effects of time. The most parsimonious among them was selected as the preferred model. This generalized (log link with Gamma distribution) model included random intercepts, linear, and quadratic slopes (supplemental equation 5). We compared mean model-predicted trajectory with observed means for each year. The trajectory approximated observed data quite well in PTSD cases and all responders, validating model accuracy (Figure 1C). We visualized advantages of the generalized versus parametric approach by plotting trajectories from the best-fitting model under both specifications for subsets of responders. The two approaches produced similar subject-specific trajectories, but those from the generalized model were less sensitive to extreme scores that deviated from an individual’s general trend (Figures S5 and S6). Finally, mixed effects models were compared to latent class growth analysis and growth mixture models, including 2-through 4-class solutions. The continuous (generalized mixed effects) model outperformed alternative models with multiple latent classes by a wide margin (Table S4). Accordingly, we reported results from the continuous model.

### Typical and Atypical Trajectories

The mean model-predicted trajectory captures the typical trend in the population (Figure 1C). Trajectory intercept (score in 2011) was 26.4 and 46.4 in the full sample and PTSD cases, respectively. In the full sample, the average change was remarkably flat with a subtle quadratic trend (inverted-U or arch-shape). This arch-shape was more pronounced, on average, in PTSD cases. Using an integrative framework for multi-level R^2^ [28], 81.9% of variance was explained in PLC scores across responders and time points; 73.9% was variance in intercepts, 8.0% was variance in slopes, whereas less than 1% was explained by the fixed effects of time, and 18.1% was residual transient fluctuations. These short-lived deviations from individual’s trajectory had SD = 5.67. For PTSD cases, 72.7% of the variance was explained, 56.6% by intercepts, 13.6% by slopes, 2.5% by the fixed effects of time, and 27.3% was residual fluctuations (SD = 8.26).

Parameter estimates (Table S5) indicate that trajectories differed between responders primarily in intercepts. Individual differences in linear and quadratic slopes after one year of follow-up were small, but after 10 years, differences in slopes became as large as differences in the intercepts. Linear slopes were only weakly related to intercepts (*r* = 0.13), whereas quadratic slopes were negatively related to both intercepts (*r* = -0.48) and linear slopes (*r* = -0.67). Accordingly, responders with higher PCL in 2011 had trajectories with a steeper arch-shape. Responders who changed rapidly initially, typically showed a decelerating rate of change, as the quadratic effect received progressively more weight over time to counteract the linear effect. Results were similar for PTSD cases, except differences in intercepts and slopes were somewhat smaller and larger, respectively. Individual differences in trajectories are visualized in Figure 2, for both the full sample and cases, using random subsamples and trajectories with extreme parameter values. Because of open enrollment, average trajectories were stratified by the year of responder’s baseline assessment as a sensitivity analysis (Figure S13).

**Figure 2.**
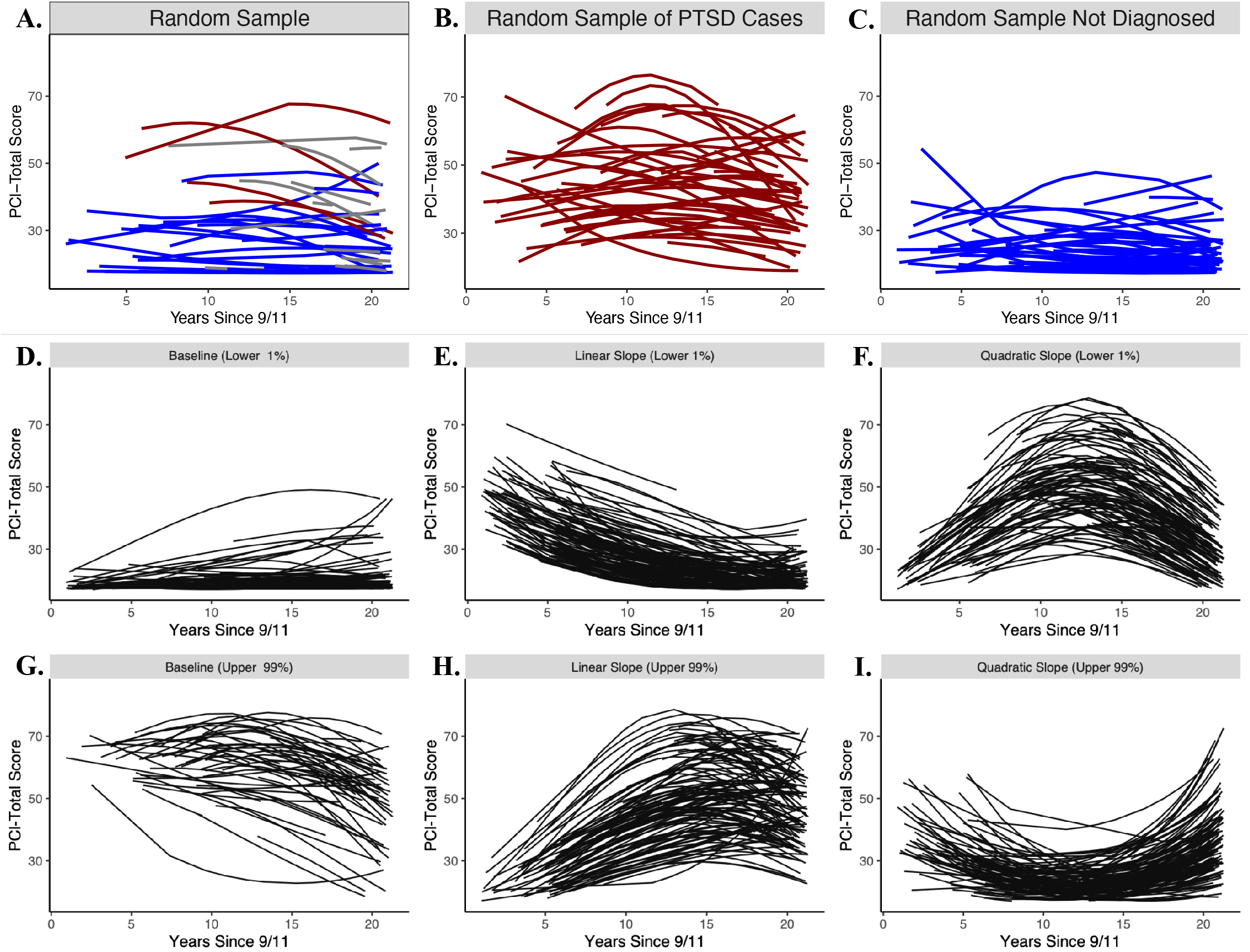
Predicted Trajectories of PTSD Symptom Severity Stratified by PTSD Diagnosis and by Trajectory Severity Quantiles **Notes**. Panels depict predicted trajectories for eight subsamples: (A) a random sample of n = 50 responders; red = diagnosis of PTSD; blue = no PTSD; gray = no diagnostic interview; (B) a random sample of n = 50 responders with a diagnosis of PTSD; (C) a random sample of n = 50 responders not diagnosed with PTSD; (D) responders with initial scores in lower 1^st^ percentile; (E) responders with linear slopes in the lower 1^st^ percentile; (F) responders with quadratic slopes in the lower 1^st^ percentile; (G) responders with initial scores in the upper 99^th^ percentile; (H) responders with linear slopes in the upper 99^th^ percentile; (I) responders with quadratic slope in the upper 99^th^ percentile. Panels D – I for responders diagnosed with PTSD are reported in Figure S10.

To further characterize variability in trajectories, we plotted change in predicted PCL since 2002 over increasing time-lags for responders with ≥3 observations (Figure 3). Change in observed PCL since 2002 over increasing time-lags can be found in the supplement (Figure S14). In responders with the lowest initial symptoms, median change was nearly zero. However, some people in this group showed substantial change. The top decile of change experienced PCL increases of ≥11 points in 20 years, whereas the bottom decile showed small decreases (∼5 points). Responders with initial symptoms in 37-46 range showed a small median improvement, decreasing by 4 points in 20 years. Change around this trend was more symmetrical with the top decile increasing ≥11 points, and the bottom decile decreasing ≥19 points. Responders with initial symptoms of 57-66 showed a more pronounced median improvement. The top decile remained at high symptom level, but nearly everyone else improved. Overall, individual differences in predicted change increased over time, typically achieving maximual variability 15-years after 9/11.

**Figure 3.**
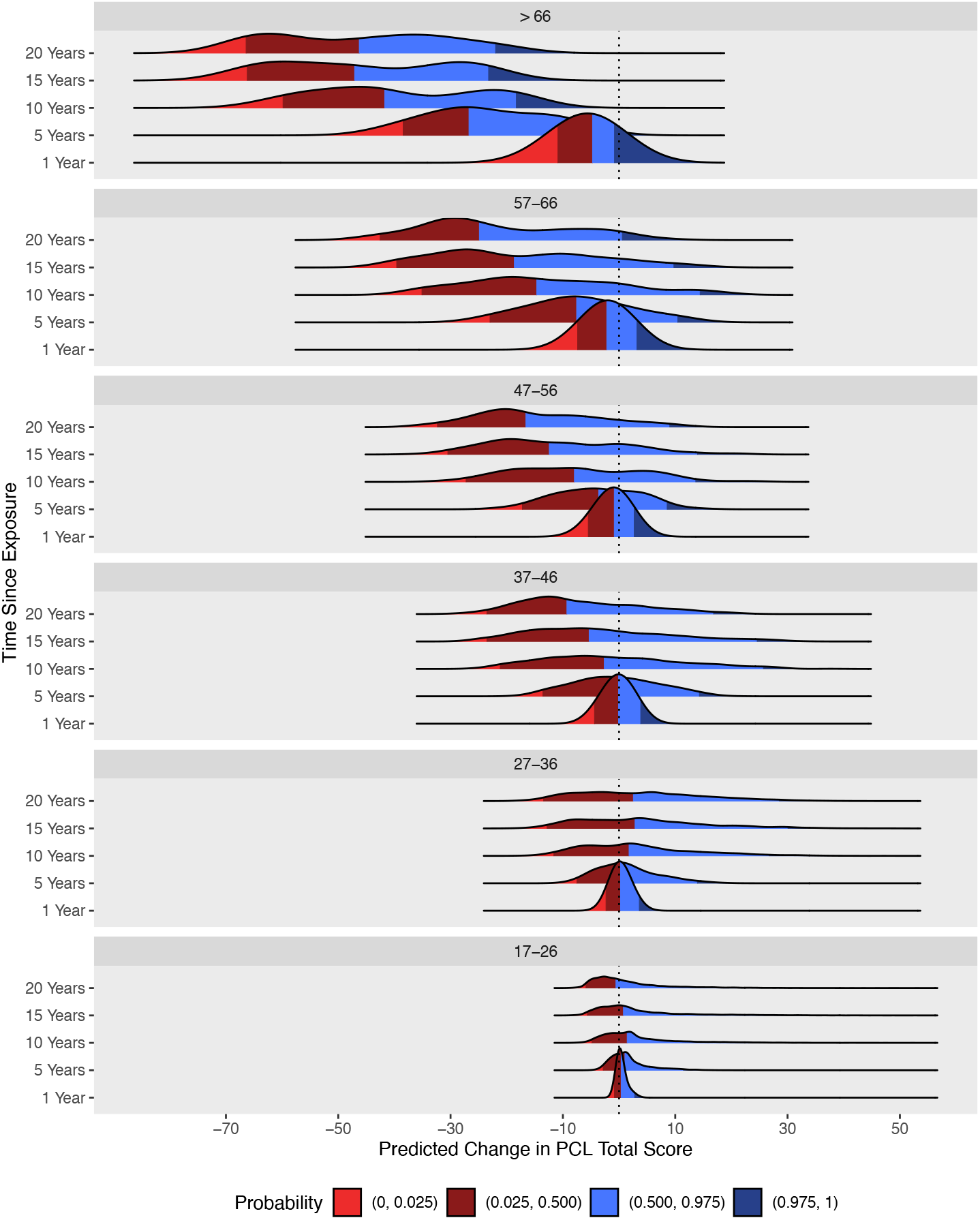
Distributions of Changes in Predicted PCL Scores Across Increasing Time Lags Stratified By PCL Score 1-Year After 9/11 **Notes**. Given the intercept-slope correlations, plots were stratfied by predicted 2002 PCL scores. Kernal density plots of predicted changes in PCL scores (x-axis) across increasing time lags (y-axis) grouped by predicted PCL scores at first visit (top label of panel). The three verital lines and colors denote the lower 2.5% tail, median, and upper 97.5% tail of each distribution. These results are expressed numerically as percentiles of predicted change in Table S5.

### Time Until Clinically Significant Change

Results are depicted in Figure 4. In the full sample, half of all responders (49%) experienced worsening symptoms over 20 years. This was even more common in PTSD cases (66%), and 50% of cases already experienced worsening after 8.60 years (95% Confidence Interval = [7.52, 10.30]). Analysis of symptom improvement was restricted to responders with intake PCL ≥ 27 to allow for a 10-point decrease. In the full sample, 50% experienced improvement over 7.00 years (95% CI = 6.61, 7.45), and 82% over 20 years. Among PTSD cases, improvement was slower, with 50% improving over 8.88 years (95% CI = 8.01, 9.79) and 76% over 20 years.

**Figure 4.**
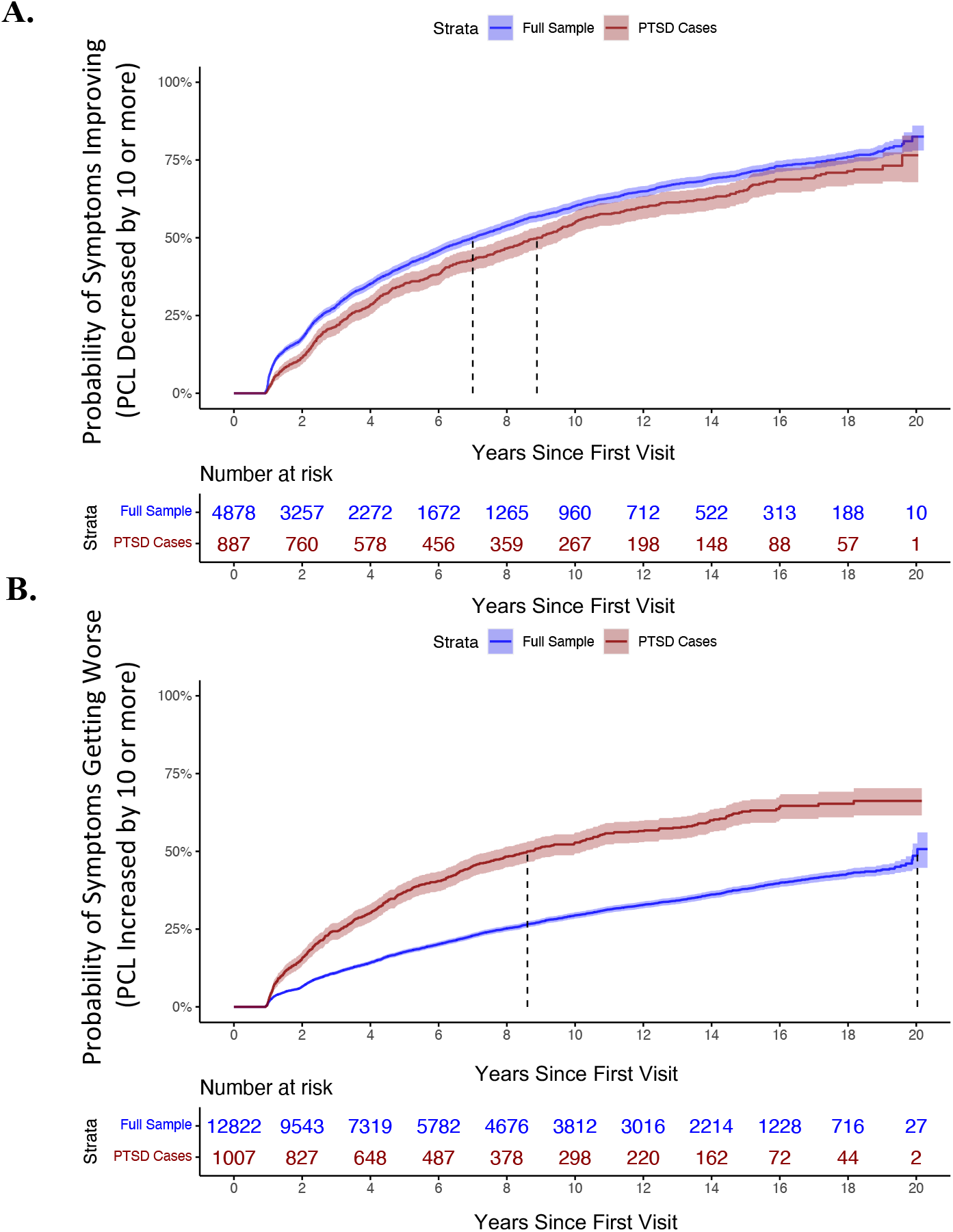
Time Until Symptoms Get Better (Panel A) or Worse (Panel B) **Notes**. The complement of Kaplan-Meier non-parametric survival probabilities are plotted with shaded regions indicating 95% confidence bands. The dashed vertical lines indicate the median survival times for the full sample and for responders with a timelife diagnosis of PTSD. Kaplan-Meier curves stratified on diagnosis of PTSD, no PTSD, and no interview (missing data) are reported in Figure S12.

## Discussion

The present study documented the 20-year prevalence, temporal stability, average and atypical trajectories, and rates of worsening and improvement of PTSD symptoms in WTC responders. It is one of the largest and longest studies of PTSD to date. Prevalence of clinically elevated symptoms ranged from 8% to 15% over two decades but has decreased in recent years. These results help explain inconsistencies across prevalence studies of PTSD observed in combat veterans [4, 5]. Symptoms changed slowly in the short-term, but retest correlations decreased to .49 after 20 years. PTSD cases showed even greater change over time. For instance, half of the cases experienced clinically significant improvement over 9 years, although significant worsening was equally common. Overall, responders differed substantially in their trajectories and ∼10% experienced a worsening or persistence of symptoms after 20 years. This reinforces conclusions of other long-term PTSD studies [5] that significant treatment needs remain decades after trauma.

Stability of PTSD symptoms in WTC responders across a few years was very similar to retest correlations of .71 to .77 found in other populations [15]. Likewise, long-term stability was on par with 19-year retest correlation in Israeli veterans (.48)[17]. Accordingly, PTSD symptoms are less stable than height, weight, and BMI [29], and about as stable as blood pressure and personality traits [29, 30]. This pattern indicates the occurrence of true change in PTSD over time, rather than just fluctuating around an individual’s set-point. Moreover, symptoms were considerably less stable in PTSD cases, with a 20-year retest correlation of only .21; PTSD cases were more likely to significantly worsen and less likely to improve than responders overall. Greater stability in the full sample is partly due to the many resilient individuals who remained asymptomatic throughout the period.

Among PTSD cases, 50% experienced improvement over seven years and 82% over 20 years, which is substantially slower than previous estimates [13]. This might be due to prior studies relying on change in diagnostic status as the primary indicator of improvement. Some participants may no longer meet PTSD criteria because of change in just one symptom, whereas we required a 10-point decrease in PCL, which is relatively stringent. Notably, almost as many cases worsened (66% over 20 years) as improved, so the population average decreased only modestly, from a PCL of 46 in 2011 to 39 in 2022.

Mean trajectory of the full sample was even flatter. However, substantial differences were observed between individual trajectories, both in initial levels and in rates of change. Differences in initial levels likely reflect risk factors such as genetic vulnerabilities and severity of WTC exposures [31], whereas rates of change probably differ due to more recent experiences. These experiences might include exposure to treatment, occupational changes (e.g., retirement), and other life events [16]. Examination of these processes is beyond the present scope, but the longitudinal model developed here provides a powerful framework for future analyses.

Differences in the rate of change were substantial. After 20 years, top and bottom deciles of change diverged by up to 44 points on PCL. Change was related to the initial level, and responders with greater symptoms in 2002 improved the most on average. However, responders in the top decile of change were very symptomatic in 2022 regardless of the initial level. They either maintained high symptoms or worsened significantly. This is consistent with the previously-reported chronic PTSD trajectory, estimated to have 10.6% prevalence across trauma-exposed populations [7]. This group is an urgent priority for health systems and public health efforts, as available resources have not been effective for them.

Another crucial finding was that after parametric assumptions were relaxed (i.e., the linear link function and Gaussian distribution of errors), a model with continuous variation in trajectories was preferred over latent class and growth mixture models. These findings are broadly consistent with a dimensional model of PTSD, which views affected individuals as experiencing extreme manifestations of what are otherwise typical and healthy expressions of natural human variation, in contrast to traditional taxonomic perspectives that emphasize discrete groups or classes of affected individuals [32]. As such, we found no evidence of discrete groups of symptom trajectories, consistent with the view that the common 4-class description of PTSD course [7] might be an artifact of fitting a parametric model to data that has substantial skew. Indeed, simulation studies have found that even slight departures from normality and linearity can lead to the over-extraction of latent classes [33, 34], even when there are no distinct classes of trajectories [35].

The observed pattern of short-term stability and long-term change has scientific and clinical implications. Studies that seek to predict or modify course of PTSD need either long follow-ups or large samples, as we found that short-term change is nearly all transient and true change accumulates slowly. Many longitudinal analyses are prone to confusing change and individual differences in PTSD levels. Mixed effects models can separate change and stable differences, but they require a minimum of three time points, and more follow-ups increase their power further. Our findings suggest that standard brief interventions are unlikely to achieve lasting change and need to be continued for extended periods of time. We observed substantial changes in some responders, but these were typically transient, with responder returning to their usual level within two years.

One limitation of the present study is that responders joined the program at different points throughout the two decades, and the number and length of follow-up assessments varied. Fortunately, we found that time of the first visit did not affect the trajectories. Also, models used in this study are flexible and do not require the same number of assessments or consistent time-lag between them. Second, diagnostic interviews were completed only on 56.9% of responders. This provided us with a sufficient sample to study trajectories of PTSD cases, but we could not examine changes in diagnostic status. Third, we have not investigated effects of treatment or other potential determinants of trajectories. These questions require detailed exposition and will be addressed in a follow-up study. Finally, WTC responders are similar to combat veterans, but they do not represent the full diversity of people with PTSD. Consequently, the generalizability of findings to other populations, especially those with more women and greater racial diversity, has to be examined.

While considering these limitations, the present study makes important and novel contributions to understanding of the long-term course of PTSD, as one of the first to examine 20-year trajectories in detail. We found no evidence of discrete trajectory groups and instead leveraged the power and precision of dimensional modeling. It revealed a non-linear pattern of mean-level change, with symptoms peaking in 2011 and gradually improving thereafter. However, responders differed substantially in their trajectories, and ∼10% experienced symptom worsening or persistence after 20 years, despite availability of free treatment. This group is a high priority for public health efforts, as significant treatment needs remain. Moreover, our findings indicate that for most responders, many years passed before a lasting change in symptoms occurred. Clinicians need to anticipate this in treatment planning. Moreover, these long-term instabilities may suggest that diagnostic efforts relying on a single time-point to screen for PTSD, especially in the period right after trauma exposure, necessarily miss a large population of individuals whose symptoms may worsen over a period of years or even decades. Concurrently, most individuals with the highest acute symptoms see gradual reductions in symptoms over time. Together, these results may imply that PTSD is a chronic condition that can last at least 20 years after initial trauma exposure and has highly variable symptom courses that improve in some individuals but can emerge or worsen for a decade prior to any improvement.

## Data Availability

Data not provided in the article because of space limitations are not publicly shared. All data and related documentation underlying the reported results will be made available after the de-identification of sensitive information and participant information. The authors will share the data with qualified investigators whose proposal to analyze the data has been approved by an internal review committee.

## References

1. Davis, L.L., et al., The economic burden of posttraumatic stress disorder in the United States from a societal perspective. The Journal of Clinical Psychiatry, 2022. 83(3): p. 40672.

2. Jordan, H.T., et al., Persistent mental and physical health impact of exposure to the September 11, 2001 World Trade Center terrorist attacks. Environmental health, 2019. 18: p. 1–16.

3. Lowell, A., et al., 9/11-related PTSD among highly exposed populations: a systematic review 15 years after the attack. Psychological Medicine, 2018. 48(4): p. 537–553.

4. Solomon, Z., et al., Predictors of PTSD trajectories following captivity: A 35-year longitudinal study. Psychiatry research, 2012. 199(3): p. 188–194.

5. Marmar, C.R., et al., Course of posttraumatic stress disorder 40 years after the Vietnam War: Findings from the National Vietnam Veterans Longitudinal Study. JAMA psychiatry, 2015. 72(9): p. 875–881.

6. Koenen, K.C., et al., Risk factors for course of posttraumatic stress disorder among Vietnam veterans: a 14-year follow-up of American Legionnaires. Journal of consulting and clinical psychology, 2003. 71(6): p. 980.

7. Galatzer-Levy, I.R., S.H. Huang, and G.A. Bonanno, Trajectories of resilience and dysfunction following potential trauma: A review and statistical evaluation. Clinical psychology review, 2018. 63: p. 41–55.

8. Able, M.L. and D.M. Benedek, Severity and symptom trajectory in combat-related PTSD: a review of the literature. Current Psychiatry Reports, 2019. 21: p. 1–8.

9. Feder, A., et al., Risk, coping and PTSD symptom trajectories in World Trade Center responders. Journal of psychiatric research, 2016. 82: p. 68–79.

10. Pietrzak, R., et al., Trajectories of PTSD risk and resilience in World Trade Center responders: an 8-year prospective cohort study. Psychological medicine, 2014. 44(01): p. 205–219.

11. Lee, D.J., et al., The 20-year course of posttraumatic stress disorder symptoms among veterans. Journal of abnormal psychology, 2020. 129(6): p. 658.

12. Rosellini, A.J., et al., Recovery from DSM-IV post-traumatic stress disorder in the WHO World Mental Health surveys. Psychological Medicine, 2018. 48(3): p. 437–450.

13. Morina, N., et al., Remission from post-traumatic stress disorder in adults: a systematic review and meta-analysis of long term outcome studies. Clinical psychology review, 2014. 34(3): p. 249–255.

14. Bui, E., et al., Course of posttraumatic stress symptoms over the 5 years following an industrial disaster: a structural equation modeling study. Journal of traumatic stress, 2010. 23(6): p. 759–766.

15. Livingston, N.A., et al., Longitudinal assessment of PTSD and illicit drug use among male and female OEF-OIF veterans. Addictive Behaviors, 2021. 118: p. 106870.

16. Zvolensky, M.J., et al., Post-disaster stressful life events and WTC-related posttraumatic stress, depressive symptoms, and overall functioning among responders to the World Trade Center disaster. Journal of psychiatric research, 2015. 61: p. 97–105.

17. Ginzburg, K., T. Ein-Dor, and Z. Solomon, Comorbidity of posttraumatic stress disorder, anxiety and depression: a 20-year longitudinal study of war veterans. Journal of affective disorders, 2010. 123(1-3): p. 249–257.

18. Pietrzak, R.H., et al., The burden of full and subsyndromal posttraumatic stress disorder among police involved in the World Trade Center rescue and recovery effort. Journal of psychiatric research, 2012. 46(7): p. 835–842.

19. Dasaro, C.R., et al., Cohort Profile: World Trade Center Health Program General Responder Cohort. Int J Epidemiol, 2017. 46(2): p. e9.

20. Bromet, E.J., et al., DSM-IV post-traumatic stress disorder among World Trade Center responders 11–13 years after the disaster of 11 September 2001 (9/11). Psychological medicine, 2016. 46(4): p. 771–783.

21. Blanchard, E.B., et al., Psychometric properties of the PTSD Checklist (PCL). Behav Res Ther, 1996. 34(8): p. 669–73.

22. Robins, L.N., et al., Diagnostic Interview Schedule for the DSM-IV (DIS-IV) Washington University School of Medicine; St. Louis, MO, 2000.

23. First, M.B., et al., Structured Clinical Interview for DSM-IV Axis I Disorders–Patient Edition (version 2). New York: Biometric Research Institute, 1995.

24. Bromet, E., et al., DSM-IV post-traumatic stress disorder among World Trade Center responders 11–13 years after the disaster of 11 September 2001 (9/11). Psychological medicine, 2016. 46(4): p. 771–783.

25. Hunter, J.E. and F.L. Schmidt, Methods of meta-analysis: Correcting error and bias in research findings. 2004: Sage.

26. Monson, C.M., et al., Change in posttraumatic stress disorder symptoms: do clinicians and patients agree? Psychological assessment, 2008. 20(2): p. 131.

27. Blanchard, E.B., et al., Psychometric properties of the PTSD Checklist (PCL). Behaviour research and therapy, 1996. 34(8): p. 669–673.

28. Rights, J.D. and S.K. Sterba, Quantifying explained variance in multilevel models: An integrative framework for defining R-squared measures. Psychological methods, 2019. 24(3): p. 309.

29. Fujita, F. and E. Diener, Life satisfaction set point: stability and change. Journal of personality and social psychology, 2005. 88(1): p. 158.

30. Mann, F.D., C.G. DeYoung, and R.F. Krueger, Patterns of cumulative continuity and maturity in personality and well-being: Evidence from a large longitudinal sample of adults. Personality and Individual Differences, 2021. 169: p. 109737.

31. Waszczuk, M.A., et al., Polygenic prediction of PTSD trajectories in 9/11 responders. Psychological Medicine, 2020: p. 1–9.

32. Hawn, S.E., et al., Conceptualizing traumatic stress and the structure of posttraumatic psychopathology through the lenses of RDoC and HiTOP. Clinical Psychology Review, 2022. 95: p. 102177.

33. Bauer, D.J. and P.J. Curran, Distributional assumptions of growth mixture models: implications for overextraction of latent trajectory classes. Psychological methods, 2003. 8(3): p. 338.

34. Bauer, D.J. and P.J. Curran, The integration of continuous and discrete latent variable models: potential problems and promising opportunities. Psychological methods, 2004. 9(1): p. 3.

35. Bauer, D.J., Observations on the use of growth mixture models in psychological research. Multivariate behavioral research, 2007. 42(4): p. 757–786.

