## supplement for "Long-Term Trajectories of Posttraumatic Stress Disorder Symptoms: A 20-Year Longitudinal Study of World Trade Center Responders"

### Supplemental Methods

#### Sample Characteristics

The sample is predominately male (*n* =11,652; ~91%), with comparatively few females (*n* = 1,170; ~9%). Race was "unknown" for a large number of patients (n = 5221; ~41%); Among patients who reported their race, the majority identified as White (n = 6,635; 52%), followed by Black (n = 438; ~3%), multi-racial (n = 413; ~3%), and Asian (n = 86; ~1%), while all other races comprised less than 1% of the sample, specifically Native American (*n* = 19; <1%), and Pacific Islander (n = 10; <1%).

#### Descriptive Statistics

Compared to a normal distribution, positive third and fourth order moments indicate that, year by year, cross-sectional distributions of PCL scores were peaked with comparatively thick and positively skewed tails, suggesting a possible floor effect. Indeed, depending on the year, 16% to 34% of responders reported few to no symptoms, indicated by PCL scores equal to 17 or 18 (on a scale that ranges from= [17 - 85]). In the full sample, the average PCL score was relatively stable from year to year (range of means = [26.04, 29.70], range of SD = [11.24, 14.77]), and there were higher average and greater variation in symptoms for responders with a lifetime diagnosis of PTSD (range of means = [38.22, 49.59], range of SD = [13.05, 17.68]).

#### Mixed Effects Models

In the equations below, $PCL_{ti}$ is the repeatedly measured PTSD symptoms at time t for responder i. $[\beta_0 + d_{i0}]$ is the intercept or predicted score for responder i when $Time_{ti} - 10 = 0$. $\beta_0$ is the fixed effect or expected sample mean for the intercept and $d_{0i}$ is the random effect or individual deviations from the expected sample mean. $[\beta_1 + d_{1i}]$ is the random linear slope or rate of change for responder i given a one-unit change in time, where time is the number of years since the 9/11 attacks. $\beta_1$ is the fixed linear slope or expected sample mean for the linear slope and $d_{i1}$ is the random linear slope, which captures individual deviations from the expected sample mean. $[\beta_2 + d_{i2}]$ and $[\beta_3 + d_{i3}]$ are the non-linear slopes or predicted rates of change for responder i given a one-unit change in a transformation of time, either quadratic (equations 4 or 5), natural log (equation 6), square root (equation 7), or cubic (equations 8 & 9). Finally, $\in_{it}$ is an individual time-specific residual score. All models were estimated using maximum likelihood and results were compared across different R packages ("nlme", "lme4", "lcmm") to ensure consistency of findings across analytic routines.

(1) $PCL_{it} = \beta_0 + d_{i0} + \in_{ti}$

(2) $PCL_{it} = \beta_0 + d_{i0} + \beta_1 \times [Time_{ti} - 10] + \in_{ti}$

(3) $PCL_{it} = [\beta_0 + d_{i0}] + [\beta_1 + d_{i1}] \times [Time_{ti} - 10] + \in_{ti}$

(4) $PCL_{it} = [\beta_0 + d_{i0}] + [\beta_1 + d_{i1}] \times [Time_{ti} - 10] + \beta_2 \times [Time_{ti} - 10]^2 + \in_{ti}$

(5) $PCL_{it} = [\beta_0 + d_{i0}] + [\beta_1 + d_{i1}] \times [Time_{ti} - 10] + [\beta_2 + d_{i2}] \times [Time_{ti} - 10]^2 + \in_{ti}$

(6) $PCL_{it} = [\beta_0 + d_{i0}] + [\beta_1 + d_{i1}] \times Time_{ti} + [\beta_2 + d_{i2}] \times \ln[Time_{ti}] + \in_{ti}$

(7) $PCL_{it} = [\beta_0 + d_{i0}] + [\beta_1 + d_{i1}] \times Time_{ti} + [\beta_2 + d_{i2}] \times \sqrt{Time_{ti}} + \in_{ti}$

(8) $PCL_{it} = [\beta_0 + d_{i0}] + [\beta_1 + d_{i1}] \times [Time_{ti} - 10] + [\beta_2 + d_{i2}] \times [Time_{ti} - 10]^2 + \beta_3 \times [Time_{ti} - 10]^3 + \in_{ti}$

(9) $PCL_{it} = [\beta_0 + d_{i0}] + [\beta_1 + d_{i1}] \times [Time_{ti} - 10] + [\beta_2 + d_{i2}] \times [Time_{ti} - 10]^2 + [\beta_3 + d_{i3}] \times [Time_{ti} - 10]^3 + \in_{ti}$

Note, the addition of a fixed cubic slope (equation 8) had negligible impact on information criteria (e.g., $\Delta < 10$), compared to a more parsimonious model with no cubic term (equation 5), but the addition of a random cubic slope (equation 9) lowered information criteria considerably (e.g., Δ BIC 850). However, less than 1% of the variance in PCL scores was accounted for by the addition of the random cubic effect ($\Delta R^2_{\text{conditional}} = .008$), and no variance was accounted for by the addition of a fixed cubic effect ($\Delta R^2_{\text{marginal}} = .000$). Moreover, the intra-class correlation (ICC = .82 vs .83) and residual standard deviation (5.6 vs 5.5) remained largely unchanged before and after the inclusion of random cubic effects.

**Table S1.** Descriptive Statistics and Estimates of Internal Consistency for PCL Across Waves of Data Collection for the Full Sample

| | Descriptive Statistics | | | | | | | | Internal Consistency | | |
|---|---|---|---|---|---|---|---|---|---|---|---|
| PCL | n | mean | median | SD | min | max | skew | kurtosis | $\alpha$ | $\omega_T$ | $\omega_H$ |
| Visit 1 | 12822 | 28.07 | 23.00 | 13.22 | 17.00 | 85.00 | 1.64 | 2.34 | 0.95 | 0.96 | 0.86 |
| Visit 2 | 11462 | 28.21 | 23.00 | 13.53 | 17.00 | 85.00 | 1.58 | 2.05 | 0.96 | 0.96 | 0.86 |
| Visit 3 | 9975 | 27.79 | 22.00 | 13.40 | 17.00 | 85.00 | 1.62 | 2.17 | 0.96 | 0.97 | 0.87 |
| Visit 4 | 8612 | 27.66 | 22.00 | 13.40 | 17.00 | 85.00 | 1.62 | 2.16 | 0.96 | 0.97 | 0.86 |
| Visit 5 | 7390 | 27.61 | 22.00 | 13.25 | 17.00 | 85.00 | 1.60 | 2.17 | 0.96 | 0.96 | 0.85 |
| Visit 6 | 6275 | 27.67 | 22.00 | 13.32 | 17.00 | 85.00 | 1.58 | 1.95 | 0.96 | 0.97 | 0.87 |
| Visit 7 | 5374 | 27.63 | 22.00 | 13.21 | 17.00 | 85.00 | 1.58 | 2.00 | 0.96 | 0.97 | 0.85 |
| Visit 8 | 4583 | 27.52 | 22.00 | 12.97 | 17.00 | 85.00 | 1.59 | 2.10 | 0.96 | 0.96 | 0.86 |
| Visit 9 | 3865 | 27.36 | 22.00 | 12.92 | 17.00 | 85.00 | 1.64 | 2.31 | 0.96 | 0.96 | 0.86 |
| Visit 10 | 3219 | 27.34 | 22.00 | 12.88 | 17.00 | 85.00 | 1.64 | 2.31 | 0.96 | 0.96 | 0.86 |
| Visit 11 | 2606 | 27.05 | 22.00 | 12.38 | 17.00 | 85.00 | 1.59 | 2.14 | 0.96 | 0.96 | 0.86 |
| Visit 12 | 2033 | 26.93 | 22.00 | 12.34 | 17.00 | 79.00 | 1.64 | 2.35 | 0.95 | 0.96 | 0.84 |
| Visit 13 | 1447 | 27.14 | 22.00 | 12.47 | 17.00 | 79.00 | 1.55 | 1.92 | 0.96 | 0.96 | 0.85 |
| Visit 14 | 931 | 26.92 | 22.00 | 12.17 | 17.00 | 81.00 | 1.60 | 2.20 | 0.95 | 0.96 | 0.83 |
| Visit 15 | 475 | 26.34 | 21.00 | 11.93 | 17.00 | 80.00 | 1.64 | 2.42 | 0.95 | 0.96 | 0.80 |
| Visit 16 | 185 | 26.66 | 21.00 | 12.49 | 17.00 | 81.00 | 1.67 | 2.63 | 0.95 | 0.96 | 0.80 |
| Visit 17 | 44 | 24.53 | 21.00 | 8.26 | 17.00 | 51.00 | 1.52 | 1.83 | 0.94 | 0.95 | 0.70 |

Descriptive Statistics and Estimates of Internal Consistency for PCL Across Waves of Data Collection for PTSD Cases

| | Descriptive Statistics | | | | | | | | Internal Consistency | | |
|---|---|---|---|---|---|---|---|---|---|---|---|
| PCL | n | mean | median | SD | min | max | skew | kurtosis | $\alpha$ | $\omega_T$ | $\omega_H$ |
| Visit 1 | 966 | 45.83 | 45.00 | 15.77 | 17.00 | 85.00 | 0.21 | -0.75 | 0.94 | 0.79 | 0.95 |
| Visit 2 | 930 | 48.65 | 48.00 | 15.99 | 17.00 | 85.00 | 0.10 | -0.70 | 0.94 | 0.81 | 0.95 |
| Visit 3 | 924 | 46.75 | 47.00 | 15.90 | 17.00 | 85.00 | 0.16 | -0.73 | 0.94 | 0.82 | 0.95 |
| Visit 4 | 885 | 46.69 | 46.00 | 15.94 | 17.00 | 85.00 | 0.10 | -0.84 | 0.94 | 0.81 | 0.95 |
| Visit 5 | 835 | 45.76 | 45.00 | 15.53 | 17.00 | 85.00 | 0.20 | -0.65 | 0.94 | 0.81 | 0.95 |
| Visit 6 | 760 | 45.46 | 45.00 | 15.56 | 17.00 | 85.00 | 0.16 | -0.69 | 0.94 | 0.81 | 0.95 |
| Visit 7 | 665 | 44.92 | 43.56 | 15.53 | 17.00 | 83.87 | 0.16 | -0.76 | 0.94 | 0.79 | 0.95 |
| Visit 8 | 588 | 44.39 | 44.00 | 15.78 | 17.00 | 82.88 | 0.22 | -0.75 | 0.94 | 0.81 | 0.95 |
| Visit 9 | 529 | 42.95 | 42.00 | 15.71 | 17.00 | 85.00 | 0.34 | -0.59 | 0.94 | 0.80 | 0.95 |
| Visit 10 | 466 | 42.46 | 41.69 | 15.42 | 17.00 | 85.00 | 0.39 | -0.49 | 0.94 | 0.81 | 0.95 |
| Visit 11 | 378 | 42.67 | 41.22 | 14.86 | 17.00 | 82.57 | 0.29 | -0.80 | 0.93 | 0.79 | 0.95 |
| Visit 12 | 308 | 40.57 | 39.00 | 14.69 | 17.00 | 79.69 | 0.45 | -0.49 | 0.93 | 0.76 | 0.95 |
| Visit 13 | 239 | 41.45 | 40.00 | 14.83 | 17.00 | 85.00 | 0.42 | -0.41 | 0.94 | 0.77 | 0.95 |
| Visit 14 | 170 | 41.45 | 39.00 | 14.84 | 17.00 | 81.00 | 0.28 | -0.61 | 0.93 | 0.73 | 0.95 |
| Visit 15 | 102 | 40.57 | 38.50 | 13.27 | 17.00 | 72.00 | 0.34 | -0.57 | 0.92 | 0.66 | 0.94 |
| Visit 16 | 42 | 39.94 | 40.00 | 12.68 | 17.00 | 63.00 | -0.03 | -1.19 | 0.90 | 0.58 | 0.94 |
| Visit 17 | 12 | 41.64 | 40.00 | 12.58 | 20.00 | 61.00 | -0.08 | -1.41 | 0.90 | 0.57 | 0.94 |

**Notes.** n = sample size. SD = standard deviation. min. = minimum observed scores. max. = maximum observed score. $\alpha$ = Cronbach's alpha. $\omega_T$ = Omega total. $\omega_H$ = Omega hierarchical.

**Table S2.** Information Criteria Comparing Linear and Generalized Mixed Effects Models of PCL Total Scores for the Full Sample

| Linear Mixed Effect Models Estimated in Full Sample | -LL | AIC | BIC | SSBIC |
|---|---|---|---|---|
| $PCL_{ti}=\beta_0+u_{0i}+\epsilon_{ti}$ | -286652.73 | 573314.10 | 573342.02 | 573363.76 |
| $PCL_{ti}=\beta_0+\beta_1 Time_{ti}+u_{0i}+\epsilon_{ti}$ | -286526.51 | 573072.21 | 573109.43 | 573130.75 |
| $PCL_{ti}=\beta_0+\beta_1 Time_{ti}+\beta_2 Time^2_{ti}+u_{0i}+\epsilon_{ti}$ | -286335.78 | 572704.80 | 572751.33 | 572768.73 |
| $PCL_{ti}=\beta_0+\beta_1 Time_{ti}+u_{0i}+u_{1i} Time_{ti}+\epsilon_{ti}$ | -284015.18 | 568052.51 | 568108.35 | 568146.97 |
| $PCL_{ti}=\beta_0+\beta_1 Time_{ti}+\beta_2 Time^2_{ti}+u_{0i}+u_{1i} Time_{ti}+\epsilon_{ti}$ | -283833.84 | 567703.81 | 567768.95 | 567803.71 |
| **$PCL_{ti}=\beta_0+\beta_1 Time_{ti}+\beta_2 Time^2_{ti}+u_{0i}+u_{1i} Time_{ti}+u_{2i} Time^2_{ti}+\epsilon_{ti}$** | **-282359.69** | **564761.02** | **564854.07** | **564913.71** |
| $PCL_{ti}=\beta_0+\beta_1 Time_{ti}+\beta_2 \log(Time)_{ti}+u_{0i}+u_{1i} Time_{ti}+u_{2i} \log(Time)_{ti}+\epsilon_{ti}$ | -282735.55 | 565502.38 | 565595.44 | 565665.44 |
| $PCL_{ti}=\beta_0+\beta_1 Time_{ti}+\beta_2 \sqrt{Time_{ti}}+u_{0i}+u_{1i} Time_{ti}+u_{2i} \sqrt{Time}_{ti}+\epsilon_{ti}$ | -282568.45 | 565167.42 | 565260.48 | 565331.23 |
| **$PCL_{ti}=\beta_0+\beta_1 Time_{ti}+\beta_2 Time^2_{ti}+\beta_3 Time^3_{ti}+u_{0i}+u_{1i} Time_{ti}+u_{2i} Time^2_{ti}+\epsilon_{ti}$** | **-282350.63** | **564760.33** | **564862.69** | **564915.04** |
| $PCL_{ti}=\beta_0+\beta_1 Time_{ti}+\beta_2 Time^2_{ti}+\beta_3 Time^3_{ti}+u_{0i}+u_{1i} Time_{ti}+u_{2i} Time^2_{ti}+u_{3i} Time^3_{ti}+\epsilon_{ti}$ | -281876.74 | 563820.29 | 563959.88 | 564044.99 |

| Generalized Mixed Effects Models Estimated in Full Sample | -LL | AIC | BIC | SSBIC |
|---|---|---|---|---|
| $PCL_{ti}=e^{(\beta 0+u0i+\epsilon ti)}$ | -255322.44 | 510650.882 | 510678.800 | 510703.183 |
| $PCL_{ti}=e^{(\beta 0+\beta 1Timeti+u0i+\epsilon ti)}$ | -255047.73 | 510103.449 | 510140.673 | 510173.185 |
| $PCL_{ti}=e^{(\beta 0+\beta 1Timeti+\beta 2Time2ti+u0i+\epsilon ti)}$ | -254928.67 | 509867.348 | 509913.877 | 509954.517 |
| $PCL_{ti}=e^{(\beta 0+\beta 1Timeti+u0i+u1iTimeti+\epsilon ti)}$ | -249294.66 | 498601.312 | 498657.147 | 498705.914 |
| $PCL_{ti}=e^{(\beta 0+\beta 1Timeti+\beta 2Time2ti+u0i+u1iTimeti+\epsilon ti)}$ | -249128.38 | 490951.104 | 498335.892 | 498392.788 |
| **$PCL_{ti}=e^{(\beta 0+\beta 1Timeti+\beta 2Time2ti+u0i+u1iTimeti+u2iTime2ti+\epsilon ti)}$** | **-245465.55** | **497166.871** | **491044.163** | **491125.441** |
| $PCL_{ti}=e^{(\beta 0+\beta 1Timeti+\beta 2log(Time)ti+u0i+u1iTimeti+u2ilog(Time)ti+\epsilon ti)}$ | -248573.44 | 496833.797 | 497259.930 | 497341.207 |
| $PCL_{ti}=e^{(\beta 0+\beta 1Timeti+\beta 2\sqrt{T}imeti+u0i+u1iTimeti+u2i\sqrt{T}ime\ ti+\epsilon ti)}$ | -248406.90 | 490934.501 | 496926.856 | 497008.133 |
| **$PCL_{ti}=e^{(\beta 0+\beta 1Timeti+\beta 2Time2ti+\beta 3Time3ti+u0i+u1iTimeti+u2iTime2ti+\epsilon ti)}$** | **-245456.25** | **486824.970** | **491036.866** | **491126.273** |
| $PCL_{ti}=e^{(\beta 0+\beta 1Timeti+\beta 2Time2ti+\beta 3Time3ti+u0i+u1iTimeti+u2iTime2ti+u3iTime3ti+\epsilon ti)}$ | -243397.49 | 510650.882 | 486964.558 | 487086.476 |

**Notes.** Time = years since 9/11 – 10. β = fixed effects. u = random effects. ϵ = residual error term. -LL = loglikelihood. AIC = Akaike Information Criteria. BIC = Bayesian Information Criteria. Sample-size adjusted Bayesian Information Criteria.

**Table S3.** Information Criteria Comparing Linear and Generalized Mixed Effects Models of PCL Total Scores for Responders with a Diagnosis of PTSD

| Linear Mixed Effect Models Estimated for PTSD Cases | -LL | AIC | BIC | SSBIC |
|---|---|---|---|---|
| $PCL_{ti}=\beta_0+u_{0i}+\epsilon_{ti}$ | -34049.40 | 68104.79 | 68126.04 | 68157.10 |
| $PCL_{ti}=\beta_0+\beta_1Time_{ti}+u_{0i}+\epsilon_{ti}$ | -33978.81 | 67971.22 | 67999.54 | 68035.35 |
| $PCL_{ti}=\beta_0+\beta_1Time_{ti}+\beta_2Time^2_{ti}+u_{0i}+\epsilon_{ti}$ | -33823.53 | 67671.79 | 67707.20 | 67744.23 |
| $PCL_{ti}=\beta_0+\beta_1Time_{ti}+u_{0i}+u_{1i}Time_{ti}+\epsilon_{ti}$ | -33617.44 | 67251.28 | 67293.78 | 67351.48 |
| $PCL_{ti}=\beta_0+\beta_1Time_{ti}+\beta_2Time^2_{ti}+u_{0i}+u_{1i}Time_{ti}+\epsilon_{ti}$ | -33468.24 | 66964.02 | 67013.60 | 67072.51 |
| $PCL_{ti}=\beta_0+\beta_1Time_{ti}+\beta_2Time^2_{ti}+u_{0i}+u_{1i}Time_{ti}+u_{2i}Time^2_{ti}+\epsilon_{ti}$ | -33303.91 | 66640.71 | 66711.53 | 66802.15 |
| $PCL_{ti}=\beta_0+\beta_1Time_{ti}+\beta_2log(Time)_{ti}+u_{0i}+u_{1i}Time_{ti}+u_{2i}log(Time)_{ti}+\epsilon_{ti}$ | -33353.88 | 66730.22 | 66801.04 | 66902.09 |
| $PCL_{ti}=\beta_0+\beta_1Time_{ti}+\beta_2\sqrt{Time_{ti}}+u_{0i}+u_{1i}Time_{ti}+u_{2i}\sqrt{Time_{ti}}+\epsilon_{ti}$ | -33325.86 | 66673.44 | 66744.27 | 66846.06 |
| $PCL_{ti}=\beta_0+\beta_1Time_{ti}+\beta_2Time^2_{ti}+\beta_3Time^3_{ti}+u_{0i}+u_{1i}Time_{ti}+u_{2i}Time^2_{ti}+\epsilon_{ti}$ | -33303.61 | 68104.79 | 66732.49 | 66820.98 |
| $PCL_{ti}=\beta_0+\beta_1Time_{ti}+\beta_2Time^2_{ti}+\beta_3Time^3_{ti}+u_{0i}+u_{1i}Time_{ti}+u_{2i}Time^2_{ti}+u_{3i}Time^3_{ti}+\epsilon_{ti}$ | -33227.72 | 67971.22 | 66616.80 | 66746.94 |

| Generalized Mixed Effects Models Estimated for PTSD Cases | -LL | AIC | BIC | SSBIC |
|---|---|---|---|---|
| $PCL_{ti}=e^{(\beta_0+u_{0i}+\epsilon_{ti})}$ | -33530.62 | 67067.24 | 67088.48 | 67119.54 |
| $PCL_{ti}=e^{(\beta_0+\beta_1Time_{ti}+u_{0i}+\epsilon_{ti})}$ | -33430.96 | 66869.91 | 66898.24 | 66939.65 |
| $PCL_{ti}=e^{(\beta_0+\beta_1Time_{ti}+\beta_2Time2_{ti}+u_{0i}+\epsilon_{ti})}$ | -33301.41 | 66612.82 | 66648.24 | 66699.99 |
| $PCL_{ti}=e^{(\beta_0+\beta_1Time_{ti}+u_{0i}+u_{1i}Time_{ti}+\epsilon_{ti})}$ | -32685.10 | 65382.19 | 65424.69 | 65486.79 |
| $PCL_{ti}=e^{(\beta_0+\beta_1Time_{ti}+\beta_2Time2_{ti}+u_{0i}+u_{1i}Time_{ti}+\epsilon_{ti})}$ | -32529.01 | 65072.03 | 65121.60 | 65194.06 |
| **$PCL_{ti}=e^{(\beta_0+\beta_1Time_{ti}+\beta_2Time2_{ti}+u_{0i}+u_{1i}Time_{ti}+u_{2i}Time2_{ti}+\epsilon_{ti})}$** | -32129.11 | 64278.23 | 64349.05 | 64452.57 |
| $PCL_{ti}=e^{(\beta_0+\beta_1Time_{ti}+\beta_2log(Time)_{ti}+u_{0i}+u_{1i}Time_{ti}+u_{2i}log(Time)_{ti}+\epsilon_{ti})}$ | -32247.85 | 64515.71 | 64586.53 | 64690.04 |
| $PCL_{ti}=e^{(\beta_0+\beta_1Time_{ti}+\beta_2\sqrt{Time_{ti}}+u_{0i}+u_{1i}Time_{ti}+u_{2i}\sqrt{Time_{ti}}+\epsilon_{ti})}$ | -32193.49 | 64406.98 | 64477.81 | 64581.32 |
| **$PCL_{ti}=e^{(\beta_0+\beta_1Time_{ti}+\beta_2Time2_{ti}+\beta_3Time3_{ti}+u_{0i}+u_{1i}Time_{ti}+u_{2i}Time2_{ti}+\epsilon_{ti})}$** | -32129.08 | 64280.17 | 64358.08 | 64471.94 |
| $PCL_{ti}=e^{(\beta_0+\beta_1Time_{ti}+\beta_2Time2_{ti}+\beta_3Time3_{ti}+u_{0i}+u_{1i}Time_{ti}+u_{2i}Time2_{ti}+u_{3i}Time3_{ti}+\epsilon_{ti})}$ | -31910.24 | 63850.48 | 63956.72 | 64111.99 |

**Notes.** Time = years since 9/11 – 10. β = fixed effects. u = random effects. ϵ = residual error term. -LL = loglikelihood. AIC = Akaike Information Criteria. BIC = Bayesian Information Criteria. Sample-size adjusted Bayesian Information Criteria. SSBIC = sample size adjusted Bayesian information criteria.

**Table S4.** Fit Statistics Comparing Generalized Mixed Effects Models to Parametric Latent Class Growth Analysis and Linear Growth Mixture Models with Different Numbers of Classes

| Parameters | Model | G | BIC | %class1 | %class2 | %class3 | %class4 |
|---|---|---|---|---|---|---|---|
| $PCL_{ti}=\beta_0+u_{0i}+\epsilon_{ti}$ | MLM | 1 | 510678.8 | 100.00 | | | |
| $PCL_{ti}=\beta_0+\beta_1 Time_{ti}+u_{0i}+\epsilon_{ti}$ | MLM | 1 | 510140.7 | 100.00 | | | |
| $PCL_{ti}=\beta_0+\beta_1 Time_{ti}+\beta_2 Time^2_{ti}+u_{0i}+\epsilon_{ti}$ | MLM | 1 | 509913.9 | 100.00 | | | |
| $PCL_{ti}=\beta_0+\beta_1 Time_{ti}+u_{0i}+u_{1i}Time_{ti}+\epsilon_{ti}$ | MLM | 1 | 498657.1 | 100.00 | | | |
| $PCL_{ti}=\beta_0+\beta_1 Time_{ti}+\beta_2 Time^2_{ti}+u_{0i}+u_{1i}Time_{ti}+\epsilon_{ti}$ | MLM | 1 | 498335.9 | 100.00 | | | |
| **$PCL_{ti}=\beta_0+\beta_1 Time_{ti}+\beta_2 Time^2_{ti}+u_{0i}+u_{1i}Time_{ti}+u_{2i}Time^2_{ti}+\epsilon_{ti}$** | MLM | **1** | **491044.2** | **100.00** | | | |
| $PCL_{ti}=\beta_0+\beta_1 Time_{ti}+\beta_2 log(Time)_{ti}+u_{0i}+u_{1i}Time_{ti}+u_{2i}log(Time)_{ti}+\epsilon_{ti}$ | MLM | 1 | 497259.9 | 100.00 | | | |
| $PCL_{ti}=\beta_0+\beta_1 Time_{ti}+\beta_2\sqrt{Time}_{ti}+u_{0i}+u_{1i}Time_{ti}+u_{2i}\sqrt{Time}_{ti}+\epsilon_{ti}$ | MLM | 1 | 496926.8 | 100.00 | | | |
| **$PCL_{ti}=\beta_0+\beta_1 Time_{ti}+\beta_2 Time^2_{ti}+ \beta_3 Time^3_{ti} +u_{0i}+u_{1i}Time_{ti}+u_{2i}Time^2_{ti}+\epsilon_{ti}$** | MLM | **1** | **491036.9** | **100.00** | | | |
| $PCL_{ti}=\beta_0+\beta_1 Time_{ti}+\beta_2 Time^2_{ti}+ \beta_3 Time^3_{ti} +u_{0i}+u_{1i}Time_{ti}+u_{2i}Time^2_{ti}+ u_{3i}Time^3_{ti} +\epsilon_{ti}$ | MLM | 1 | 486964.6 | 100.00 | | | |
| **$PCL_{ti}=\beta_0+\beta_1 Time_{ti}+\beta_2 Time^2_{ti}+u_{0i}+u_{1i}Time_{ti}+u_{2i}Time^2_{ti}+\epsilon_{ti}$** | LCGA | 2 | 559709.1 | 14.33 | 85.67 | | |
| **$PCL_{ti}=\beta_0+\beta_1 Time_{ti}+\beta_2 Time^2_{ti}+u_{0i}+u_{1i}Time_{ti}+u_{2i}Time^2_{ti}+\epsilon_{ti}$** | GMM | 2 | 549845.6 | 39.24 | 60.75 | | |
| **$PCL_{ti}=\beta_0+\beta_1 Time_{ti}+\beta_2 Time^2_{ti}+u_{0i}+u_{1i}Time_{ti}+u_{2i}Time^2_{ti}+\epsilon_{ti}$** | LCGA | 3 | 559746.9 | 14.98 | 0.00 | 85.02 | |
| **$PCL_{ti}=\beta_0+\beta_1 Time_{ti}+\beta_2 Time^2_{ti}+u_{0i}+u_{1i}Time_{ti}+u_{2i}Time^2_{ti}+\epsilon_{ti}$** | GMM | 3 | 547307.8 | 23.51 | 51.24 | 25.25 | |
| **$PCL_{ti}=\beta_0+\beta_1 Time_{ti}+\beta_2 Time^2_{ti}+u_{0i}+u_{1i}Time_{ti}+u_{2i}Time^2_{ti}+\epsilon_{ti}$** | LCGA | 4 | 559784.8 | 14.41 | 85.59 | 0.00 | 0.00 |
| **$PCL_{ti}=\beta_0+\beta_1 Time_{ti}+\beta_2 Time^2_{ti}+u_{0i}+u_{1i}Time_{ti}+u_{2i}Time^2_{ti}+\epsilon_{ti}$** | GMM | 4 | 546850.3 | 16.91 | 14.82 | 49.79 | 18.48 |

Notes. MLM = multi-level model. LCGA = latent class growth analysis. GMM = growth mixture model. G = number of groups or classes. BIC = Bayesian Information Criteria.

**Table S5.** Results of Generalized Mixed Effects Models with a Gamma Distribution and Log Link for the Full Sample and PTSD cases

Equation: $PCL_{ti} = e^{([\beta_0 + d_{0i}] + [\beta_1 + d_{1i}] \times [Time_{ti} - 10] + [\beta_2 + d_{2i}] \times [Time_{ti} - 10]^2 + \epsilon_{ti})}$

| Parameters | Fixed Effects | | | Parameters | Correlations Between Random Effects | | |
|---|---|---|---|---|---|---|---|
| | Regression Coefficient (β) | $e^{(\beta)}$ | p | | Intercept ($d_0$) | Linear Slope ($d_1$) | Quadratic Slope ($d_2$) |
| **Full sample** | | | | | | | |
| Intercept ($\beta_0$) | 3.2730 (0.0029) | 26.3835 | < 2e-16 | Intercept ($d_0$) | 0.2484 | | |
| Linear Slope ($\beta_1$) | 0.0019 (0.0004) | 1.0018 | 2.49e-05 | Linear Slope ($d_1$) | 0.13 | 0.0264 | |
| Quadratic Slope ($\beta_2$) | -0.0009 (0.0000) | 0.9991 | < 2e-16 | Quadratic Slope ($d_2$) | -0.48 | -0.67 | 0.0032 |
| **PTSD cases** | | | | | | | |
| Intercept ($\beta_0$) | 3.8373 (0.0076) | 46.4015 | < 2e-16 | Intercept ($d_0$) | 0.1979 | | |
| Linear Slope ($\beta_1$) | 0.0064 (0.0015) | 1.0064 | 1.33e-05 | Linear Slope ($d_1$) | 0.00 | 0.0305 | |
| Quadratic Slope ($\beta_2$) | -0.0022 (0.0002) | 0.9977 | < 2e-16 | Quadratic Slope ($d_2$) | -0.39 | -0.67 | 0.0034 |

Note. β = Unstandardized coefficients are reported with standard errors in parentheses. $e^{(\beta)}$ = exponentiated coefficients. Standard deviations of random effects are reported on the diagonal of the correlation matrix of random effects. The exponentiated unstandardized intercept is the predicted PCL score 10-years after 9/11, when linear and quadratic effects equal zero. The exponentiated slopes are interpreted as average percent change in PCL scores as a linear and quadratic function of time, such that the effect of the linear coefficient remains constant over time, while the effect of the quadratic coefficient increases.

**Table S6.** Percentiles of Predicted Change in PCL-17 Total Scores Across Increasing Time Lags

| PCL at Baseline (range = 17-26) | | | | | | | | | | | | | | | |
|---|---|---|---|---|---|---|---|---|---|---|---|---|---|---|---|
| Time Lag | 1% | 5% | 10% | 20% | 25% | 30% | 40% | 50% | 60% | 70% | 75% | 80% | 90% | 95% | 99% |
| 1 Year | -1 | -1 | -1 | 0 | 0 | 0 | 0 | 0 | 0 | 1 | 1 | 1 | 2 | 2 | 3 |
| 5 Years | -4 | -3 | -2 | -1 | -1 | -1 | 0 | 0 | 1 | 2 | 3 | 4 | 7 | 10 | 14 |
| 10 Years | -6 | -5 | -4 | -3 | -2 | -1 | 0 | 1 | 2 | 4 | 5 | 7 | 13 | 19 | 29 |
| 15 Years | -7 | -5 | -4 | -3 | -3 | -2 | -1 | 0 | 2 | 4 | 5 | 8 | 15 | 21 | 33 |
| 20 Years | -7 | -6 | -5 | -4 | -3 | -3 | -2 | -1 | 1 | 2 | 3 | 5 | 11 | 17 | 28 |

| PCL at Baseline (range = 27-36) | | | | | | | | | | | | | | | |
|---|---|---|---|---|---|---|---|---|---|---|---|---|---|---|---|
| Time Lag | 1% | 5% | 10% | 20% | 25% | 30% | 40% | 50% | 60% | 70% | 75% | 80% | 90% | 95% | 99% |
| 1 Year | -3 | -2 | -2 | -1 | -1 | -1 | 0 | 0 | 1 | 1 | 1 | 2 | 2 | 3 | 4 |
| 5 Years | -9 | -7 | -6 | -4 | -4 | -3 | -1 | 0 | 2 | 4 | 5 | 6 | 10 | 12 | 16 |
| 10 Years | -14 | -11 | -9 | -7 | -6 | -5 | -3 | 0 | 3 | 7 | 8 | 12 | 18 | 23 | 33 |
| 15 Years | -15 | -12 | -10 | -8 | -7 | -6 | -3 | 0 | 3 | 7 | 10 | 12 | 20 | 26 | 35 |
| 20 Years | -15 | -12 | -11 | -9 | -7 | -6 | -3 | -1 | 2 | 6 | 8 | 11 | 18 | 23 | 31 |

| PCL at Baseline (range = 37-46) | | | | | | | | | | | | | | | |
|---|---|---|---|---|---|---|---|---|---|---|---|---|---|---|---|
| Time Lag | 1% | 5% | 10% | 20% | 25% | 30% | 40% | 50% | 60% | 70% | 75% | 80% | 90% | 95% | 99% |
| 1 Year | -5 | -4 | -4 | -2 | -2 | -1 | -1 | 0 | 0 | 1 | 1 | 2 | 3 | 3 | 4 |
| 5 Years | -15 | -13 | -11 | -7 | -6 | -5 | -3 | -1 | 1 | 3 | 5 | 7 | 10 | 13 | 16 |
| 10 Years | -22 | -20 | -17 | -13 | -11 | -10 | -6 | -3 | 1 | 5 | 7 | 10 | 18 | 23 | 28 |
| 15 Years | -25 | -21 | -18 | -15 | -14 | -12 | -8 | -5 | -1 | 3 | 6 | 8 | 16 | 24 | 28 |
| 20 Years | -25 | -22 | -19 | -16 | -15 | -13 | -11 | -8 | -4 | 1 | 3 | 4 | 11 | 14 | 20 |

| PCL at Baseline (range = 47-56) | | | | | | | | | | | | | | | |
|---|---|---|---|---|---|---|---|---|---|---|---|---|---|---|---|
| Time Lag | 1% | 5% | 10% | 20% | 25% | 30% | 40% | 50% | 60% | 70% | 75% | 80% | 90% | 95% | 99% |
| 1 Year | -6 | -5 | -4 | -3 | -3 | -2 | -1 | -1 | 0 | 1 | 1 | 1 | 2 | 2 | 4 |
| 5 Years | -19 | -16 | -13 | -10 | -9 | -8 | -5 | -3 | -1 | 2 | 3 | 4 | 6 | 7 | 14 |
| 10 Years | -27 | -24 | -22 | -18 | -16 | -14 | -11 | -7 | -3 | 2 | 3 | 5 | 9 | 11 | 23 |
| 15 Years | -32 | -28 | -25 | -22 | -20 | -19 | -15 | -10 | -7 | -2 | 0 | 2 | 6 | 12 | 21 |
| 20 Years | -36 | -30 | -27 | -24 | -22 | -20 | -18 | -13 | -10 | -6 | -5 | -2 | 4 | 8 | 11 |

| PCL at Baseline (range = 57-66) | | | | | | | | | | | | | | | |
|---|---|---|---|---|---|---|---|---|---|---|---|---|---|---|---|
| Time Lag | 1% | 5% | 10% | 20% | 25% | 30% | 40% | 50% | 60% | 70% | 75% | 80% | 90% | 95% | 99% |
| 1 Year | -8 | -8 | -6 | -5 | -4 | -4 | -2 | -2 | -1 | 0 | 0 | 0 | 2 | 3 | 3 |
| 5 Years | -25 | -23 | -19 | -15 | -12 | -12 | -9 | -7 | -5 | -1 | -1 | 0 | 7 | 9 | 11 |
| 10 Years | -36 | -35 | -32 | -25 | -22 | -21 | -18 | -15 | -9 | -4 | -3 | -1 | 11 | 14 | 18 |
| 15 Years | -41 | -38 | -37 | -31 | -29 | -27 | -25 | -18 | -13 | -8 | -6 | -3 | 2 | 9 | 14 |
| 20 Years | -45 | -43 | -38 | -31 | -30 | -29 | -28 | -24 | -18 | -11 | -9 | -7 | -1 | 0 | 7 |

| PCL at Baseline (>66) | | | | | | | | | | | | | | | |
|---|---|---|---|---|---|---|---|---|---|---|---|---|---|---|---|
| Time Lag | 1% | 5% | 10% | 20% | 25% | 30% | 40% | 50% | 60% | 70% | 75% | 80% | 90% | 95% | 99% |
| 1 Year | -12 | -9 | -9 | -9 | -9 | -8 | -5 | -4 | -4 | -3 | -3 | -2 | 0 | 1 | 1 |
| 5 Years | -39 | -39 | -37 | -31 | -29 | -28 | -27 | -25 | -19 | -16 | -14 | -11 | -9 | 1 | 1 |
| 10 Years | -62 | -62 | -57 | -54 | -53 | -48 | -45 | -42 | -31 | -25 | -25 | -19 | -19 | -2 | -2 |
| 15 Years | -68 | -68 | -64 | -63 | -60 | -58 | -54 | -45 | -35 | -28 | -27 | -27 | -22 | -10 | -10 |
| 20 Years | -67 | -67 | -66 | -63 | -63 | -63 | -58 | -42 | -35 | -33 | -31 | -24 | -22 | -21 | -21 |

**Figure S1.** Observed Trajectories of PTSD Symptoms for WTC Responders

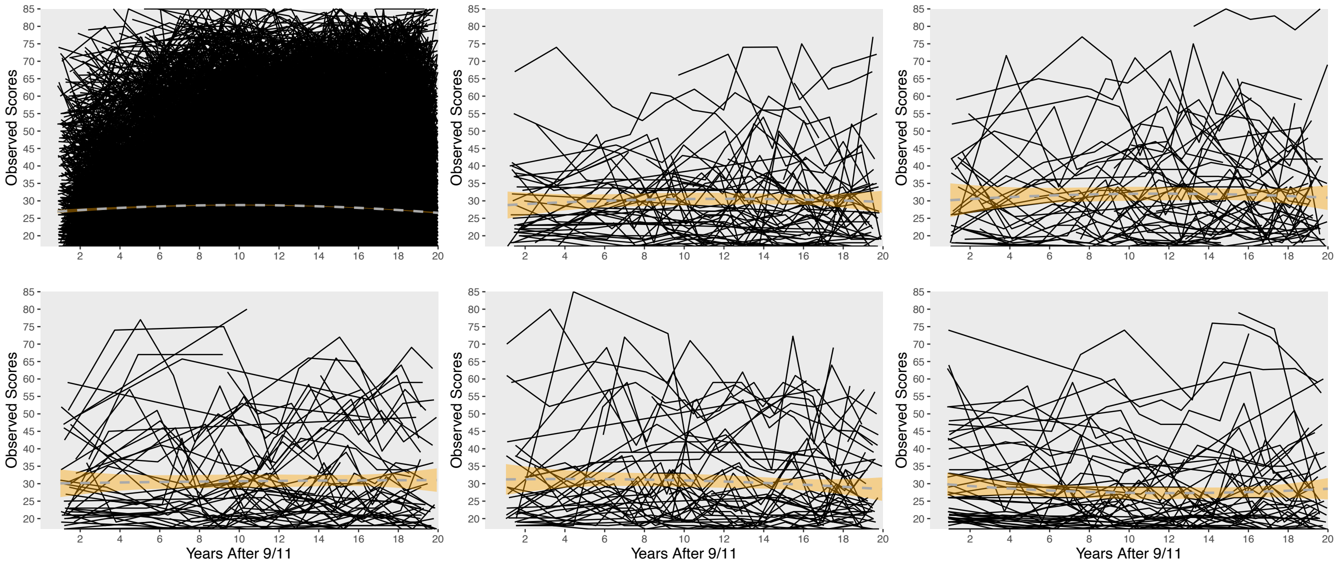


**Notes.** Top left panel, individual trajectories are plotted for the full sample. The remaining panels plot random subsamples of $n$ = 500 observations without replacement with dashed gray lines indicated quadratic trends with orange regions denoting 95% confidence bands.

**Figure S2.** Observed Trajectories of PTSD Symptoms for WTC Responders with a Diagnosis of PTSD

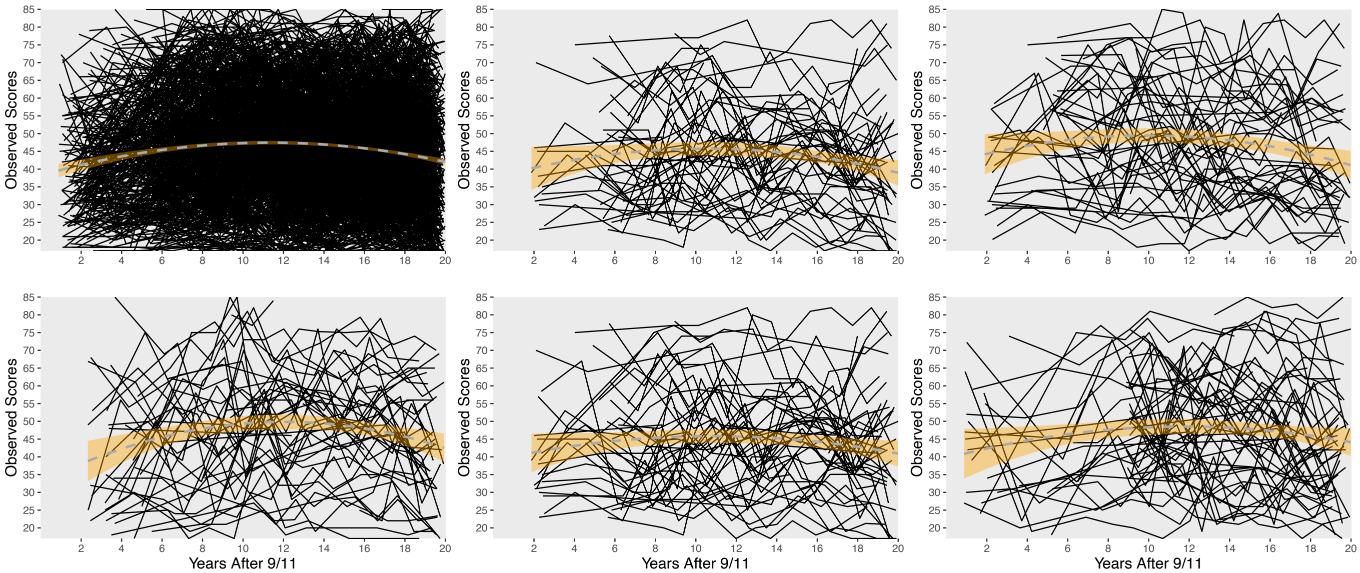


**Notes.** Top left panel, individual trajectories are plotted for all responders with a lifetime clinical diagnosis of PTSD. The remaining panels plot random subsamples of n = 500 observations without replacement. Dashed gray lines depict the average quadratic trend for each subsample with 95% confidence bands shaded in orange.

**Figure S3.** Test-Retest Correlations for PCL from 2002 to 2022 for the Full Sample

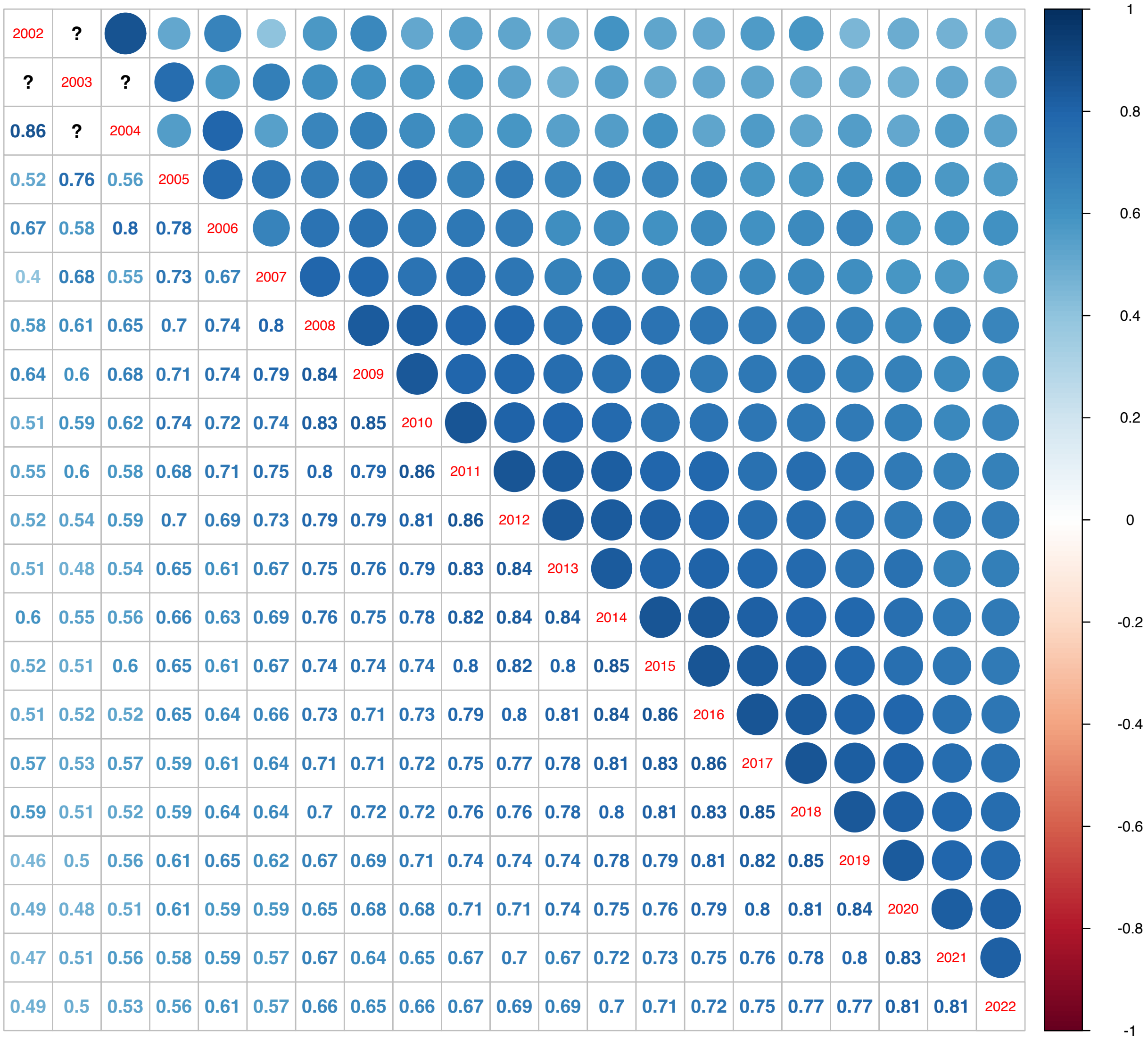


**Notes**. Pearson's product-moment correlations are reported. All correlations are statistically significant at $p < .001$. ? indicate insufficient sample sizes to estimate retest correlations.

**Figure S4.** Test-Retest Correlations for PCL from 2002 to 2022 for PTSD cases

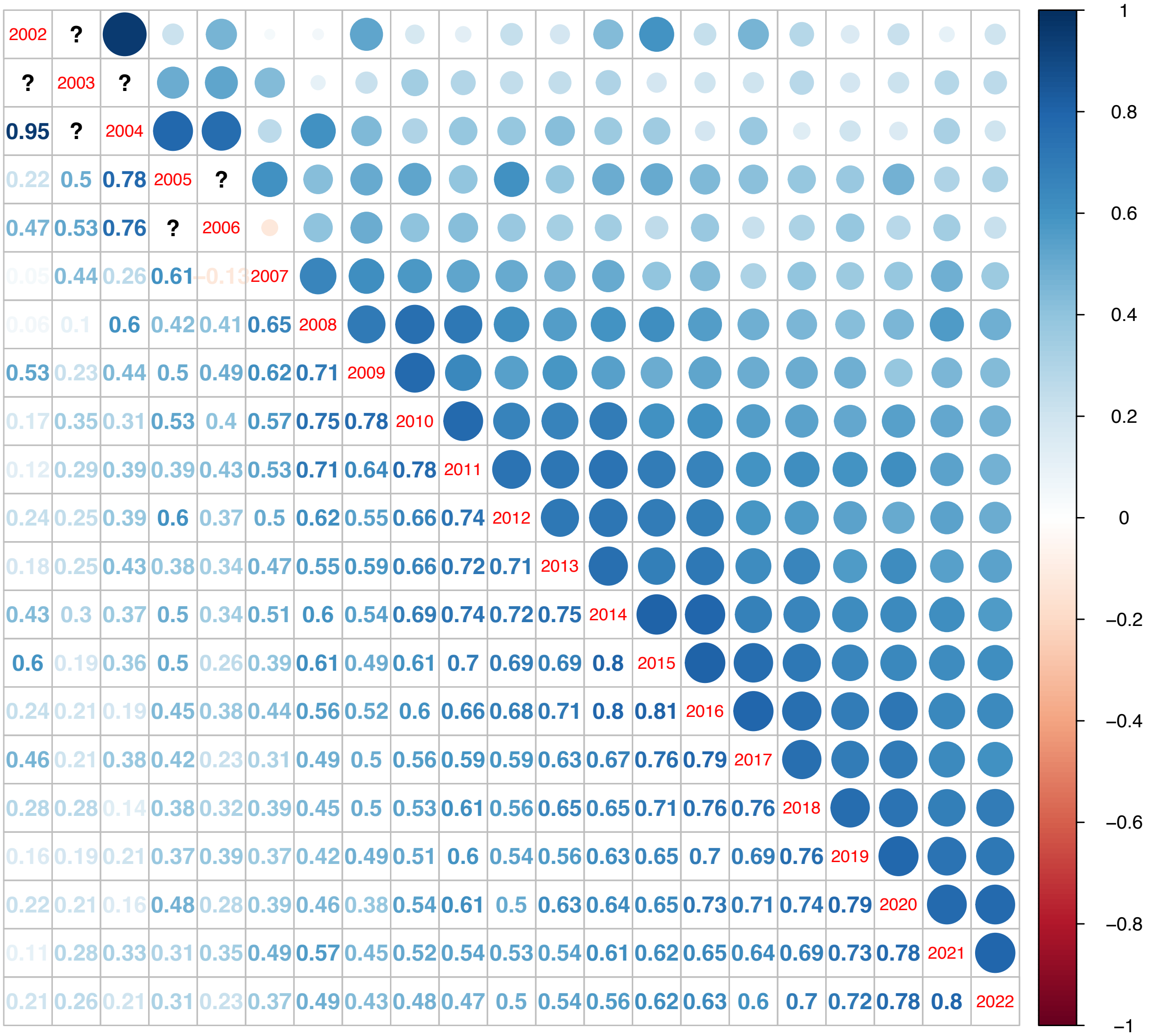


**Notes.** Pearson's product-moment correlations are reported. All correlations are statistically significant at $p < .001$. ? indicate insufficient sample sizes to estimate retest correlations

**Figure S5.** Observed Scores and Predicted Trajectories for 20 Randomly Selected Responders with Baseline Assessments after 2009

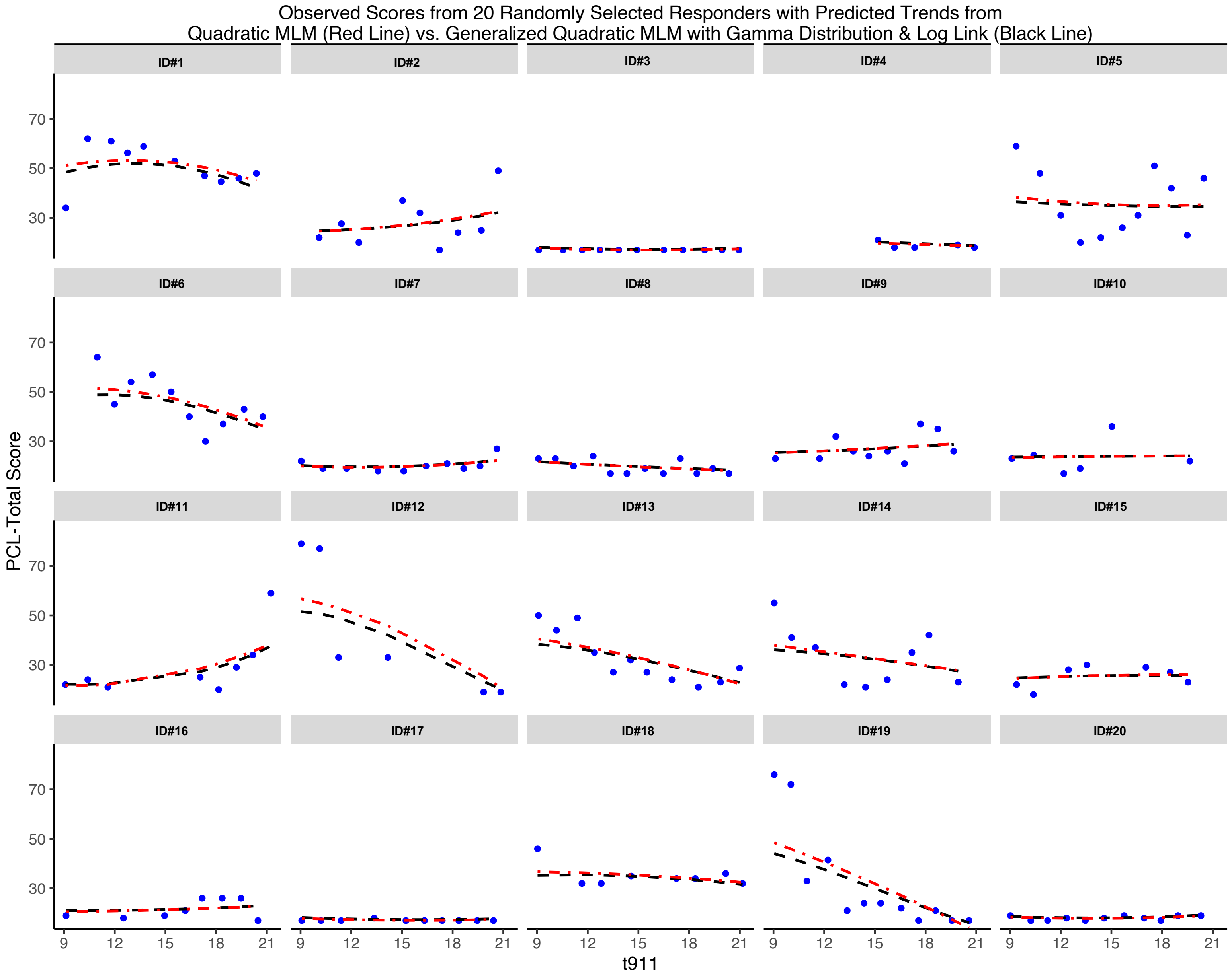

**Figure S6.** Observed Scores and Predicted Trajectories for Individual Responders Diagnosed with PTSD

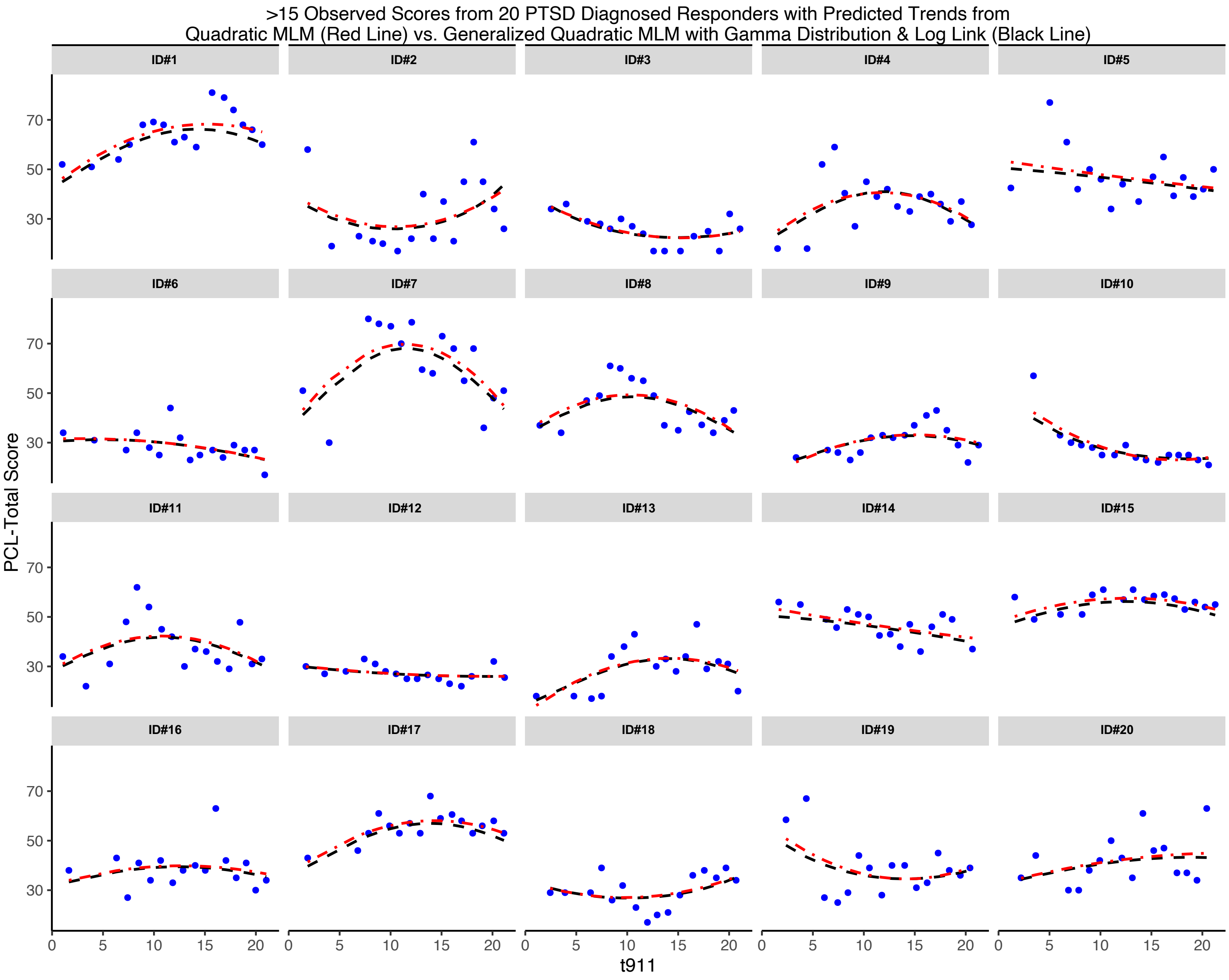

**Figure S7.** Prevalence of Clinically Elevated PTSD Symptoms (PCL ≥ 44) Stratified by Clinical Status Based on Clinical Diagnostic Interviews

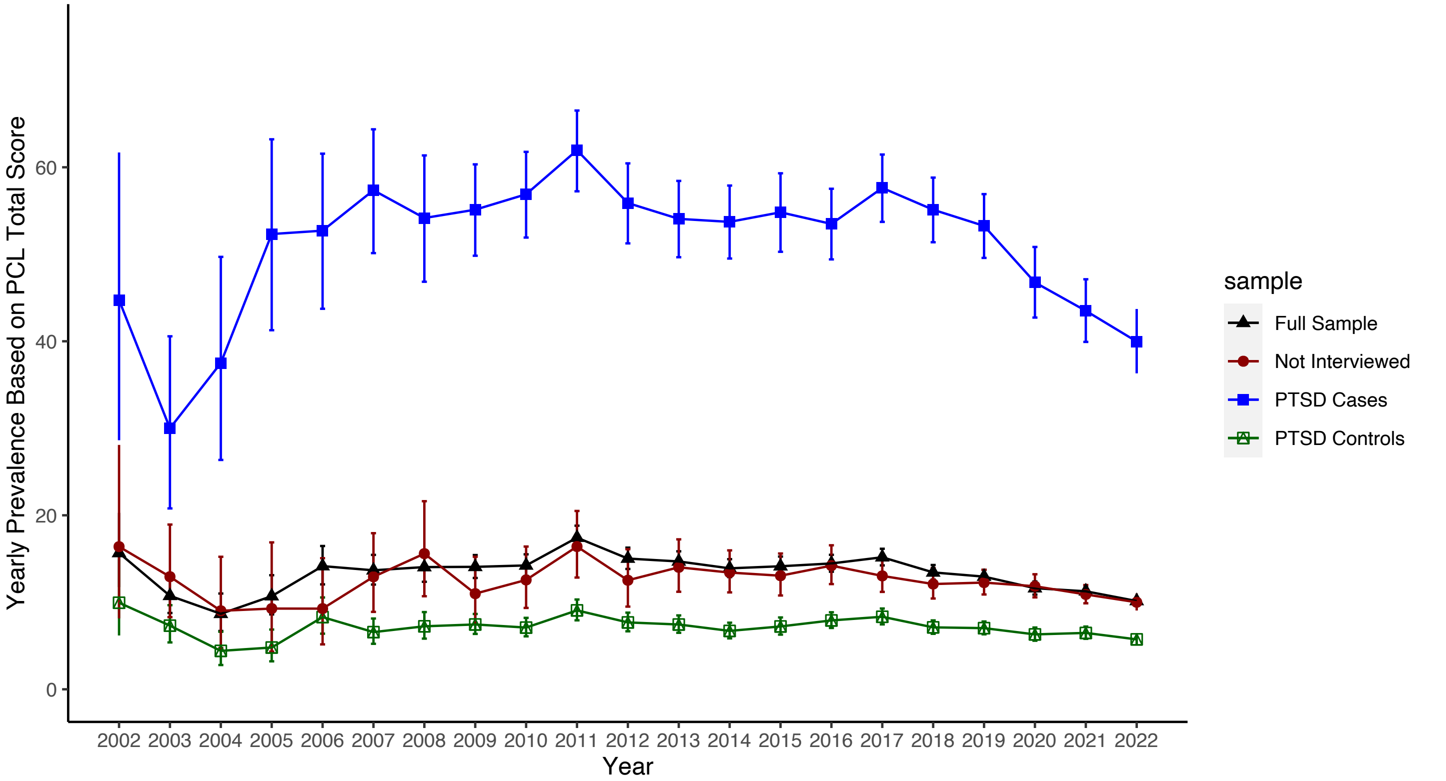


Notes. Estimated prevalence of clinically elevated symptoms (≥44 PCL-17 total score) per 100 responders with bars denoting 95% confidence intervals; subgroups diagnosed via clinical interview 10 to 15 years after 9/11.

**Figure S8.** Longitudinal Stability of PTSD Symptoms

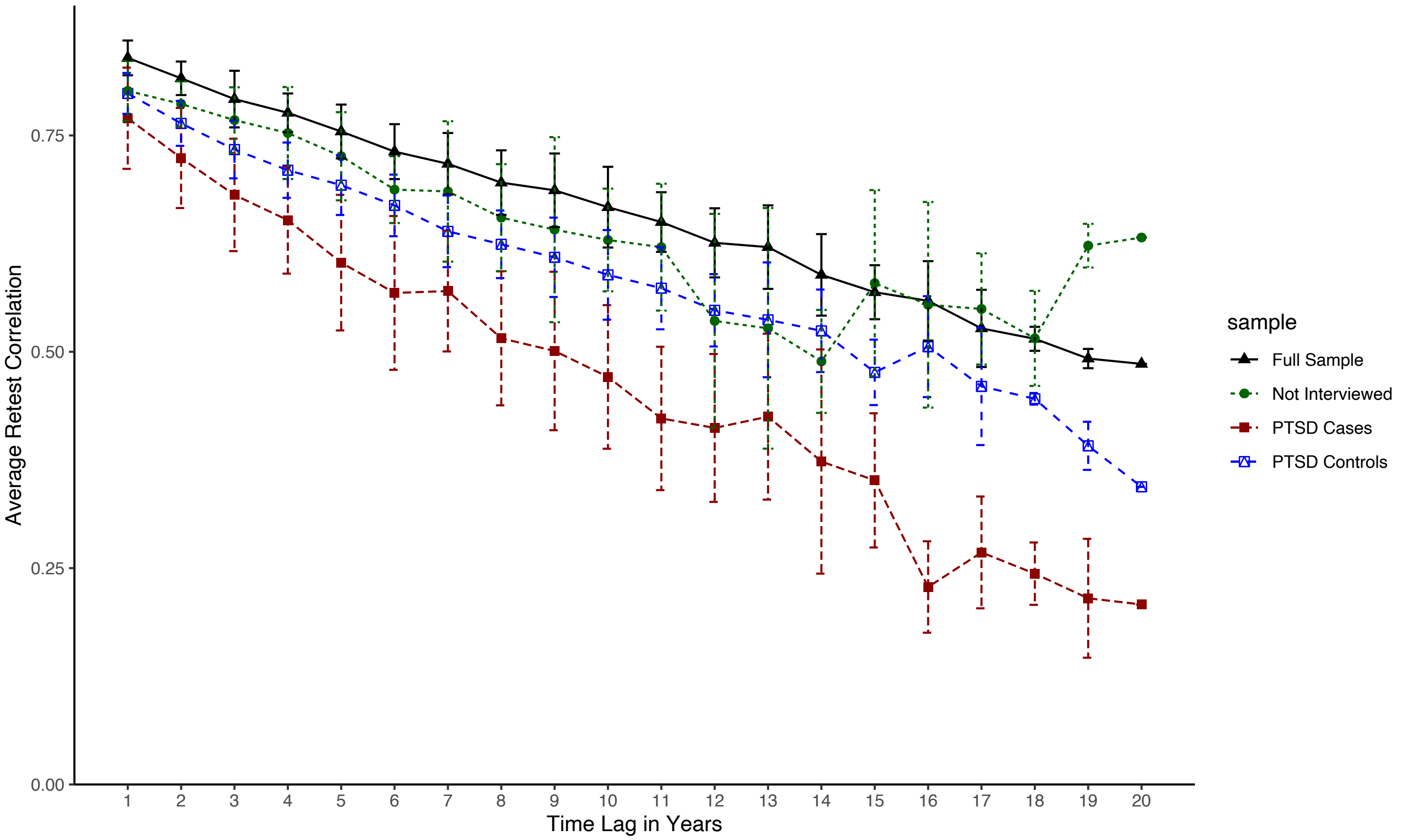


**Notes.** PTSD symptoms were measured using PCL-17 total scores. Average Pearson's correlations are reported (y-axis) across increasing time lags (x-axis) for the full sample (plotted using black triangles) and for WTC responders with a lifetime diagnosis of PTSD (plotted using red circles) with error bars denoting plus and minus 1 standard deviation.

**Figure S9.** Average Predicted Trajectory of PTSD Symptoms for the Full Sample and PTSD Cases

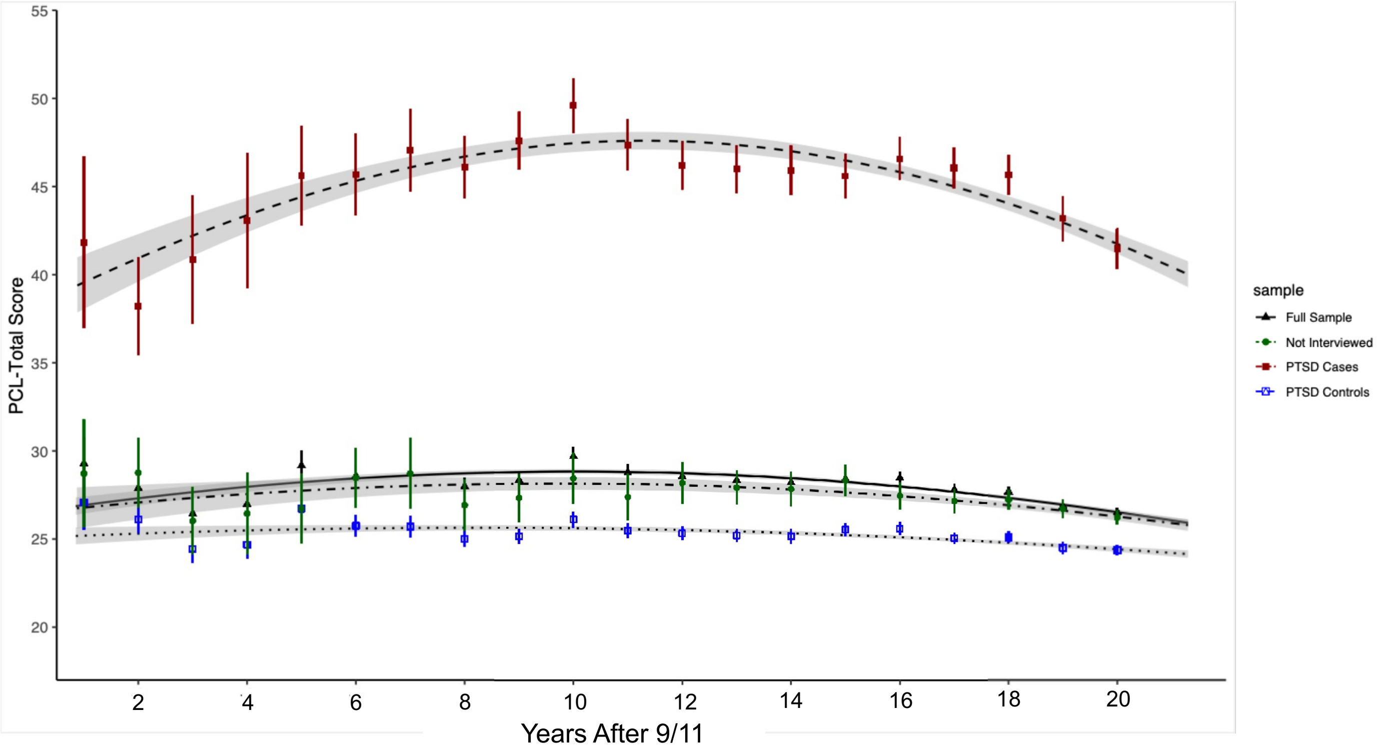


Notes. "X"s and vertical bars denote yearly observed means for the full sample (printed in blue) and for patients with a lifetime diagnosis of PTSD (printed in red) +/- 2 times the standard error of the mean. Dashed lines denote the average trajectories from generalized mixed effects models with a gamma distribution and log link with 95% confidence bands shaded in gray.

**Figure S10.** Predicted Trajectories for PTSD Cases at the Lower and Upper Extremes of Growth Parameters

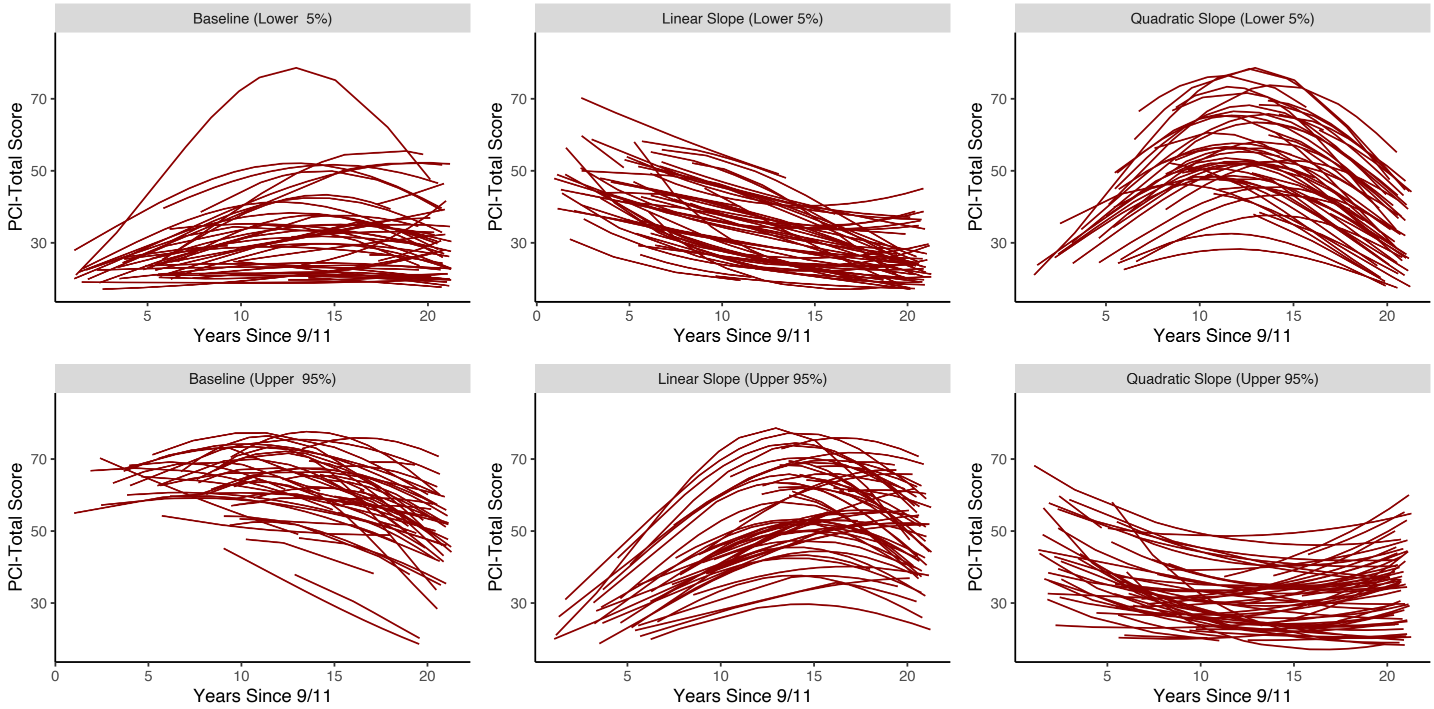


Notes. In the top two panels, predicted trajectories for responders with random intercepts, linear slopes, and quadratic slopes in the upper 95th and lower 5th percentiles are plotted on the left, middle, right panels, respectively. In the bottom panel, predicted symptom trajectories for responders in the 95th percentile of the average absolute value of the standardized random effect (right panel).

**Figure S11.** Predicted Trajectories at the Lower, Upper, and Combinatorial Extremes of Growth Model Parameters

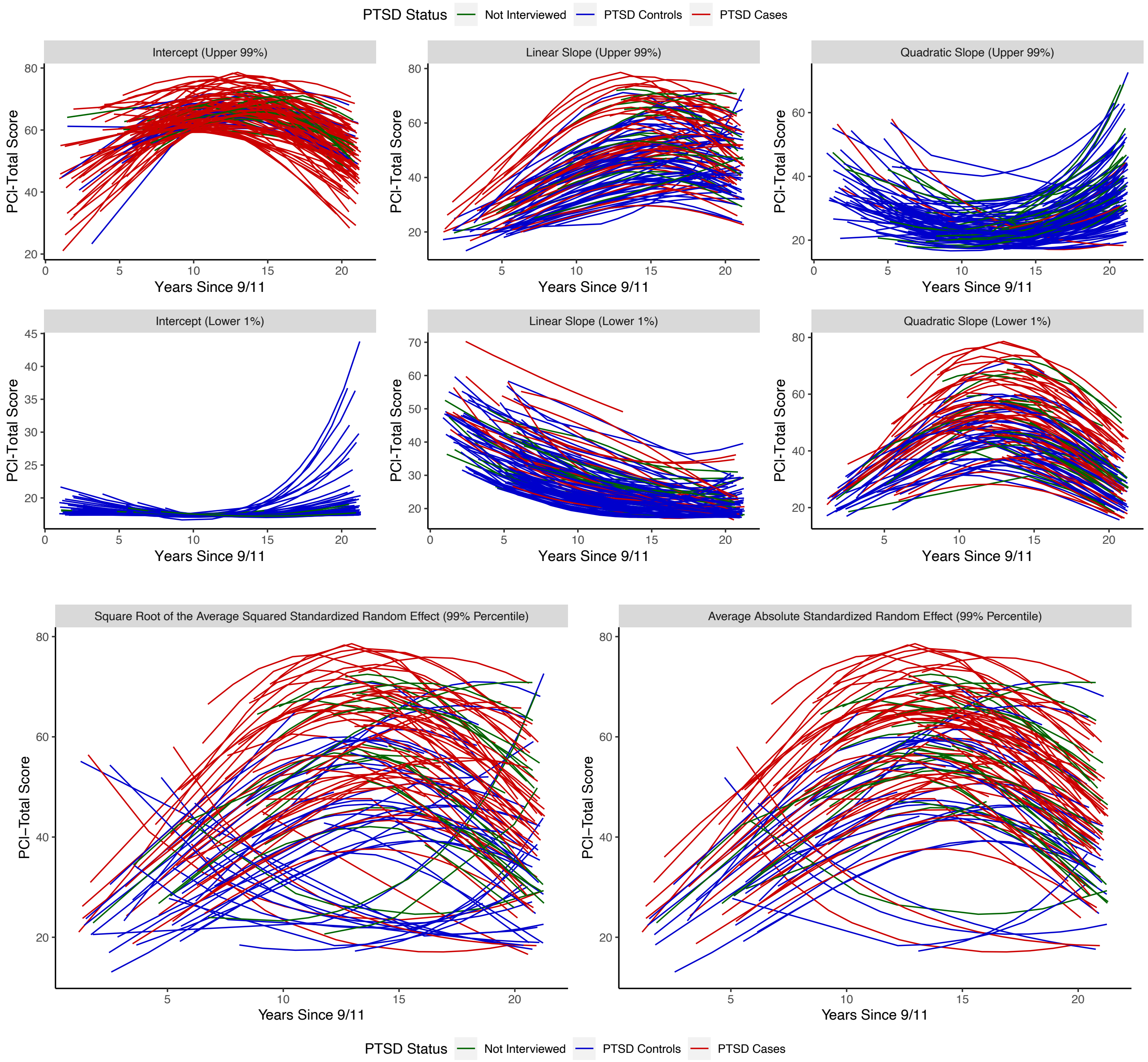


Notes. In the top two panels, predicted trajectories for responders with random intercepts, linear slopes, and quadratic slopes in the upper 95th and lower 5th percentiles are plotted on the left, middle, right panels, respectively. In the bottom panel, predicted symptom trajectories for responders in the 99th percentile of the average squared standardized random effect (left panel) and average absolute value of the standardized random effect (right panel).

**Figure S12.** Time Until Symptoms Get Better and Worse

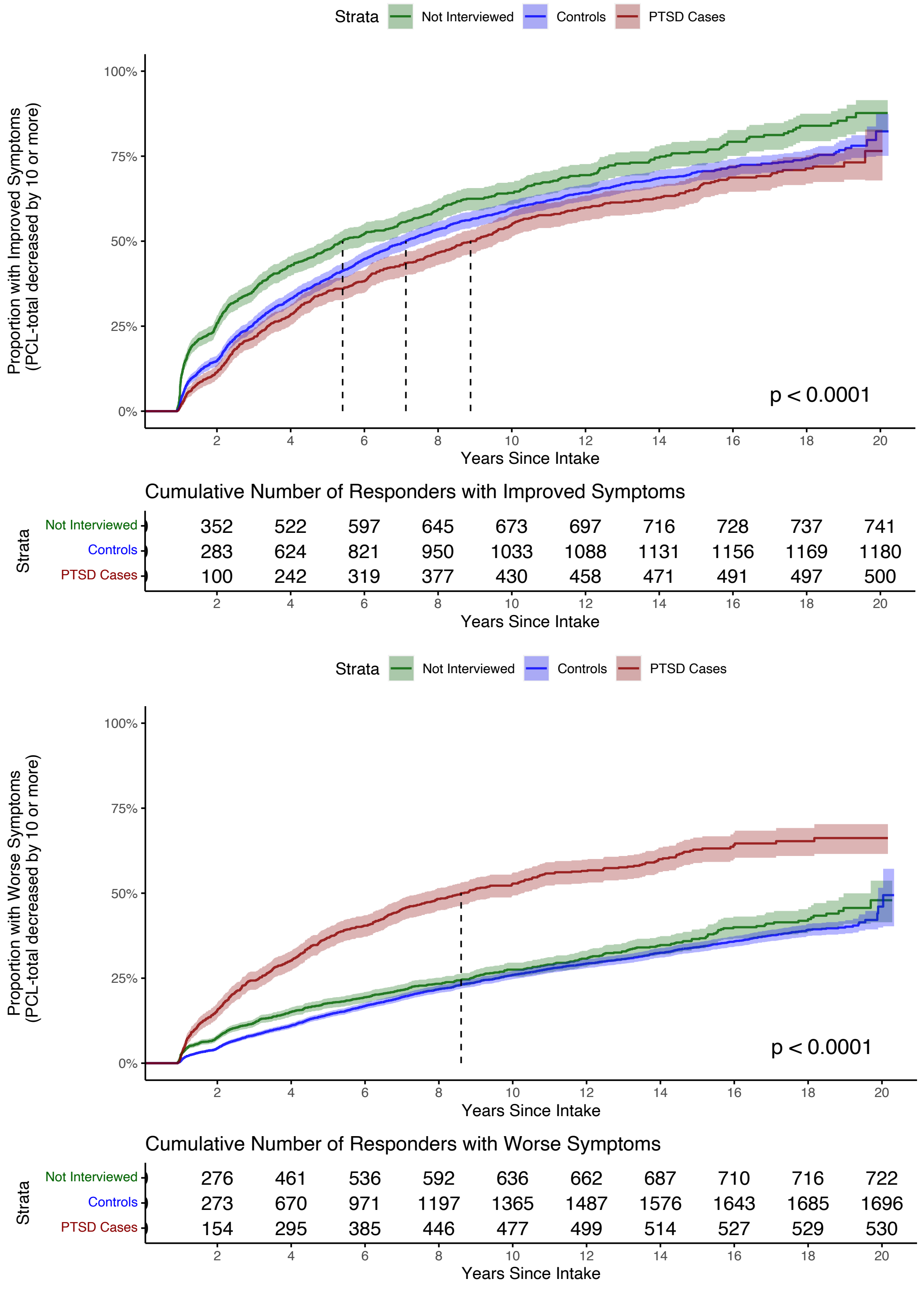


Cumulative Number of Responders with Improved Symptoms

| Strata | 2 | 4 | 6 | 8 | 10 | 12 | 14 | 16 | 18 | 20 |
|---|---|---|---|---|---|---|---|---|---|---|
| Not Interviewed | 352 | 522 | 597 | 645 | 673 | 697 | 716 | 728 | 737 | 741 |
| Controls | 283 | 624 | 821 | 950 | 1033 | 1088 | 1131 | 1156 | 1169 | 1180 |
| PTSD Cases | 100 | 242 | 319 | 377 | 430 | 458 | 471 | 491 | 497 | 500 |

Years Since Intake

Cumulative Number of Responders with Worse Symptoms

| Strata | 2 | 4 | 6 | 8 | 10 | 12 | 14 | 16 | 18 | 20 |
|---|---|---|---|---|---|---|---|---|---|---|
| Not Interviewed | 276 | 461 | 536 | 592 | 636 | 662 | 687 | 710 | 716 | 722 |
| Controls | 273 | 670 | 971 | 1197 | 1365 | 1487 | 1576 | 1643 | 1685 | 1696 |
| PTSD Cases | 154 | 295 | 385 | 446 | 477 | 499 | 514 | 527 | 529 | 530 |

Years Since Intake

**Notes.** The complement of Kaplan-Meier non-parametric survival probabilities are plotted with shaded regions indicating 95% confidence bands. The dashed lines indicate median survival times.

**Figure S13.** Average Predicted Trend in PCL Scores from the Preferred Generalized Mixed Effect Model Stratified by Baseline Year

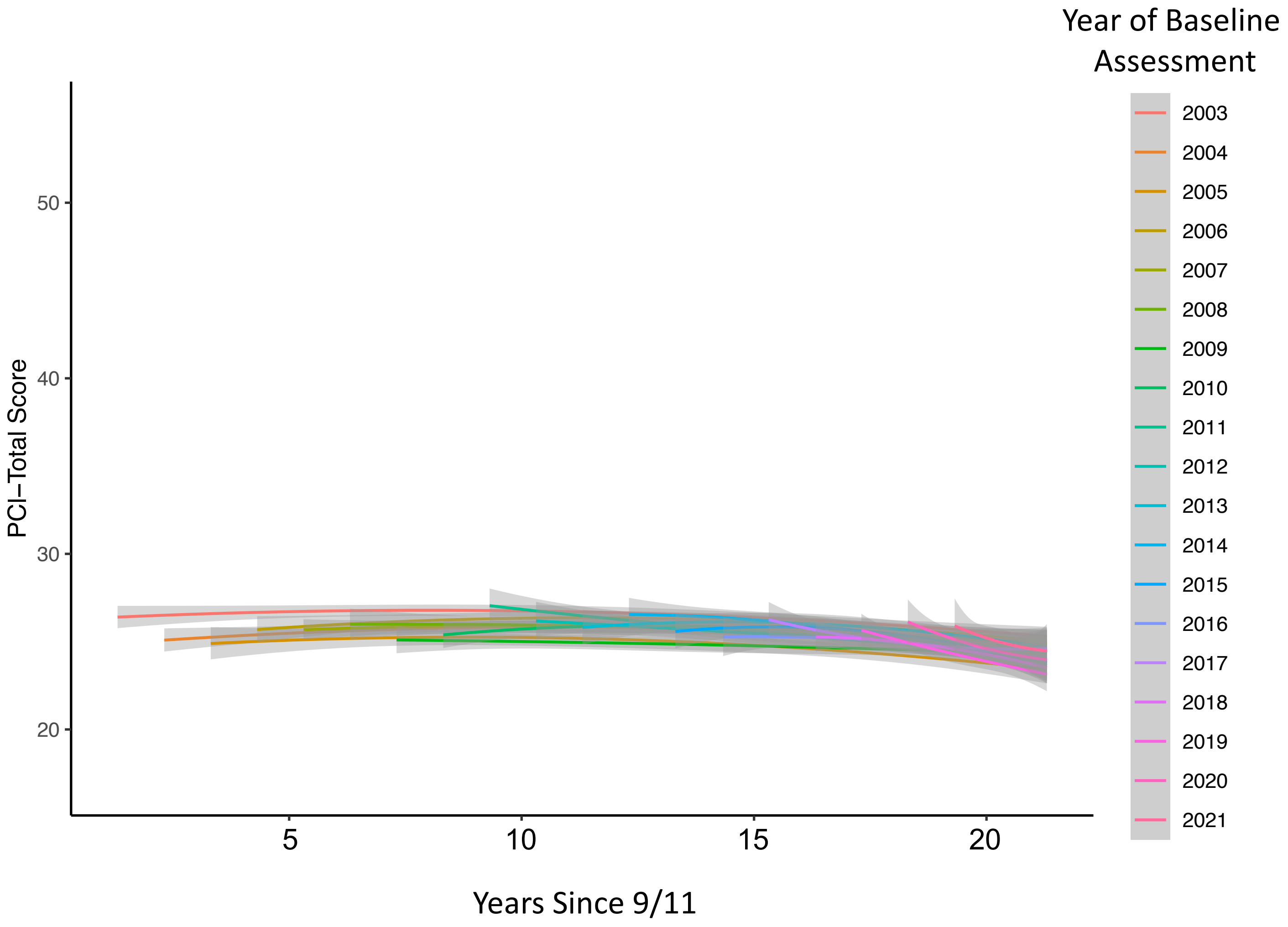


**Notes.** Each line depicts the average predicted trend for responders who completed their baseline assessment at a given year. Shaded regions depicted 95% confidence bands.

**Figure S14.** Distributions of Changes in Observed PCL Scores Across Increasing Time Lags Stratified By PCL Score 1-Year After Intake

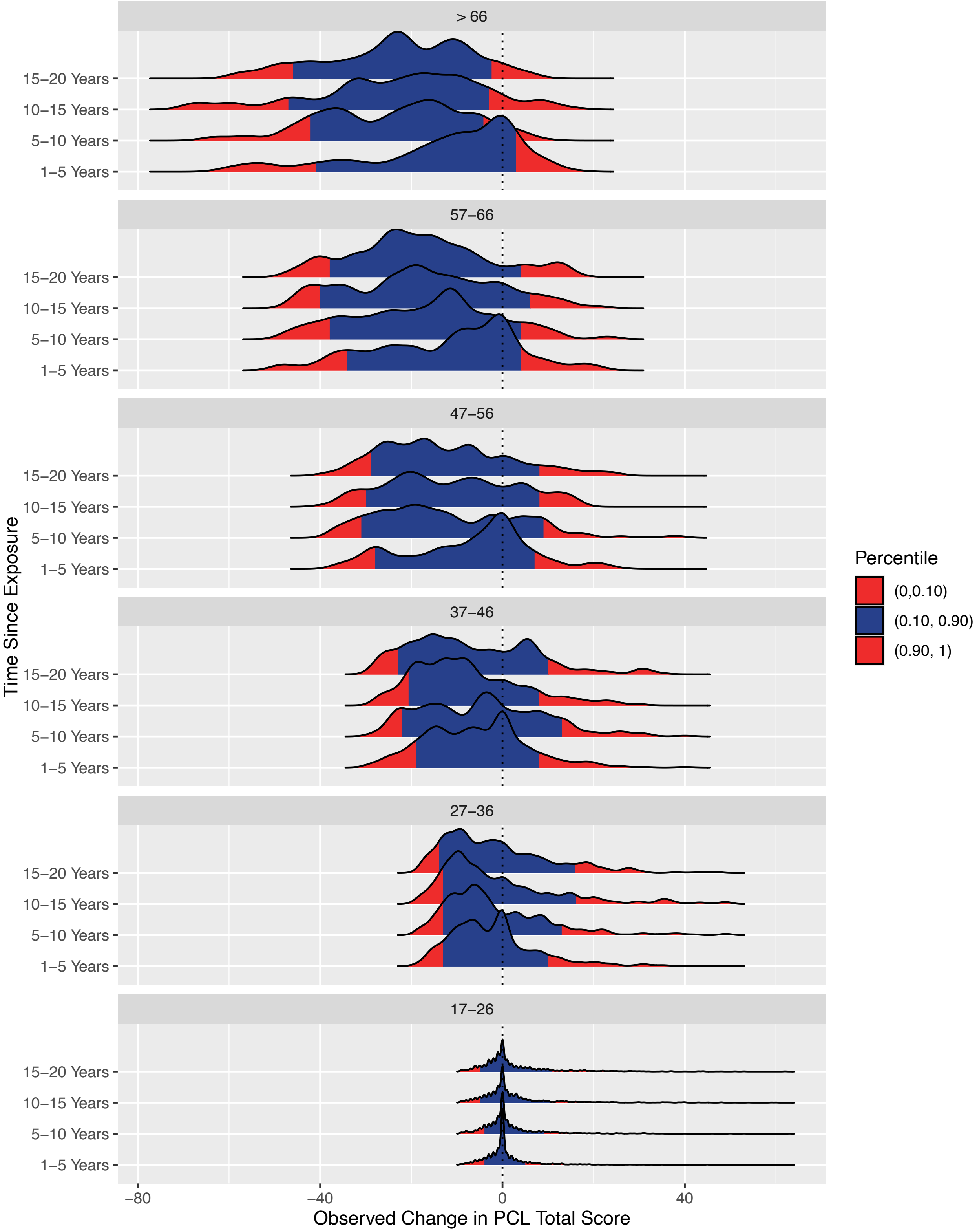


**Notes.** Given the intercept-slope correlations, plots were stratfied by observed PCl scores at in-take. Kernal density plots of observed changes in PCL scores (x-axis) across increasing time lags (y-axis) grouped by observed PCL scores at first visit (top label of panel).
